# The factor structure of attention-deficit/hyperactivity disorder in schoolchildren

**DOI:** 10.1101/2020.06.15.20126789

**Authors:** Trine Wigh Arildskov, Anne Virring, Rikke Lambek, Anders Helles Carlsen, Edmund J.S. Sonuga-Barke, Søren D. Østergaard, Per Hove Thomsen

## Abstract

This study investigated the factor structure of attention-deficit/hyperactivity disorder (ADHD) by comparing the fit of a single-factor model, a correlated model with two or three factors, and a bifactor model with one general and two or three specific factors. Different three-factor solutions that varied with regard to the specification of the item “talks excessively” as impulsivity or hyperactivity were also tested. Parent ratings on the ADHD-Rating Scale (ADHD-RS-IV) were collected in a sample of 2044 schoolchildren (1st to 3rd grade) from the general population and in a clinical sample of 165 children and adolescents with ADHD referred to a public regional child and adolescent psychiatric hospital. Confirmatory factor analyses found a satisfactory fit for most models in both samples. However, a correlated three-factor model where “talks excessively” was included as an indicator of impulsivity and especially the bifactor version of this model with one general and three specific factors fit the data slightly better in the general population. In the clinical sample, a number of models performed equally well (the same version of the correlated three-factor model and all the bifactor models). Overall, the factor structure of ADHD seems to be better characterized by a bifactor model with a strong general factor and two or three weaker specific factors. Due to the strong general factor, we suggest emphasizing the ADHD-RS-IV total score rather than the subscale scores in clinical practice.

## INTRODUCTION

Attention-deficit/hyperactivity disorder (ADHD) is a neurodevelopmental disorder defined by the presence of inattention, hyperactivity, and impulsivity across settings and to a degree that impairs daily functioning [1, 2]. The understanding of how these symptom dimensions are organized has changed considerably as the diagnostic systems have evolved over time [3]. ADHD was initially conceptualized as having three separate symptom dimensions of inattention, hyperactivity, and impulsivity in the third edition of the Diagnostic and Statistical Manual of Mental Disorders (DSM-III), but as a unidimensional construct in DSM-III-R. In both DSM-IV and DSM-5, ADHD is specified as having two symptom dimensions of inattention and combined hyperactivity/impulsivity [3]. In contrast, the tenth edition of the WHO’s International Classification of Diseases (ICD-10) [1] specify ADHD (Hyperkinetic Disorder) as having a three-factor structure. The factor structure underlying ADHD has far-reaching implications not only for the diagnostic conceptualization and assessment of the disorder (e.g., using one or more scores to determine the presence of the disorder), but also for studies of etiology, risk factors, and epidemiology.

These nosological and diagnostic developments have been based on factorial studies of the underlying structure of ADHD symptoms. Studies have tested correlated, non-hierarchical factor models typically consisting of two or three factors where symptom items are specified to load on only one of these factors. While some studies of the correlated factor models have suggested that the three-factor model has the best fit [4–7], the two-factor structure of ADHD has generally been supported and preferred by many because of the strong interrelatedness of hyperactivity and impulsivity [8–13]. More recently, bifactor models [14] specifying a general ADHD factor accounting for the shared variance between all 18 symptom items, and two or three specific factors accounting for the unique residual variance within the factor not accounted for by (and independent of) the general factor (e.g., [15–17]), are gaining support [16]. In these models, each symptom loads on the general factor *and* on one of the specific factors, and the correlations between all specified factors are restricted to zero (i.e., orthogonality) [14, 16]. Studies have quite consistently supported bifactor models of ADHD over traditional correlated models across diverse ages, informants, measures, and samples (e.g., [18–26]).

Uncertainty remains as to whether hyperactivity and impulsivity are better represented as one or two (specific) factors (e.g., [22]). Most studies support a bifactor model with two specific factors [16], although quite a few studies failed to compare this model with a bifactor model with three specific factors [9, 22, 27–31]. Of those that have, some studies have found the bifactor model with three specific factors to have a better fit in young children [21, 32] and adults [24, 25, 33]. Another issue yet to be resolved is where the item “talks excessively” should be placed. Some studies have included it as an indicator of hyperactivity (e.g., [6, 21, 22, 34, 35]), others of impulsivity (e.g., [24, 25, 32, 33, 36]), reflecting differences between DSM-IV/DSM-5 and ICD-10, respectively. Crucially, the placement of this item may explain the mixed findings in the literature regarding the structure of impulsiveness and hyperactivity and the overall factor solution favored. To the best of our knowledge, no published study with children or adolescents has addressed the role of the “talks excessively” symptom in determining the factor structure of ADHD. Finally, studies of bifactor models have been criticized for relying almost exclusively on traditional fit statistics to determine model fit [37–39] and relatively few studies have examined the bifactor models of ADHD using additional psychometric indices [16]. Such indices are important to address, especially in light of the weak and poorly-defined specific Hyperactivity/impulsivity factor found after these indices were estimated in a recent review of 18 bifactor studies of ADHD [16].

This study had two objectives. First, to explore if the factor structure of ADHD is better represented by a single-factor model, a correlated multi-factor model (with two or three factors), or a bifactor model (with one general and two or three specific factors) in a large general population sample of young schoolchildren and in a clinical sample of children and adolescents with ADHD from a previous study [40]. In this context, we also tested whether “talks excessively” is better conceptualized as an impulsivity (ICD-10) or as a hyperactivity symptom (DSM) by comparing different three-(bi)factor models. Second, to establish Danish norms for the *ADHD-Rating Scale IV Home version* (ADHD-RS-IV) [41, 42] using a much larger general population sample than has previously been possible for the age range of 6–9 [43]. Although the ADHD-RS-IV is the only questionnaire targeting all ADHD symptoms in children/adolescents that has been validated in Denmark [43], the available Danish norms do not include percentiles and were based on only 74 to 123 parent reports across the different gender by age group combinations emphasizing the need for updated norms in a larger sample.

## METHODS

### General population sample

An electronic survey was conducted to obtain a representative sample of Danish schoolchildren attending 1st, 2nd, and 3rd grade. The survey was conducted in two phases with responses collected from May 2016 to August 2017 in the Aarhus Municipality, Denmark (municipal population of 331,332 people by the second quarter of 2016 [44]). A flow chart illustrating the inclusion of participants is available in Online Resource 1.

In phase I, the survey invitation was distributed through an existing web-based intranet facility used for communication between home and school to parents of children attending 1st, 2nd, and 3rd grade at public schools in the municipality. The Department for Children and Young People provided written information about the survey in a weekly newsletter to school principals and vice principals of all 46 public schools in the municipality. Next, all schools were contacted, out of which 21 schools participated (five schools declined, and 20 schools did not reply). An invitation letter to parents was then either posted on the web-based intranet (n=20) or handed out (n=1).

In phase II, the invitation was distributed via an online secure digital mailbox (“e-Boks”) linked to the Danish personal registration number to parents of children aged 7, 8, and 9 (by February 1, 2017) who resided in Aarhus Municipality. Only one parent/caregiver (randomly chosen if more than one parent had the same registered address as the target child(ren)) received the invitation to participate. A total of 9286 parents of 9831 children received the invitation (222 parents of 243 children did not receive the invitation as they did not have “e-Boks”). The majority of parents had one child in the age group (94.2 %) followed by two and three children (5.8 %). The invitation encouraged parents to complete the survey for all of their children within the designated age range and grade level.

Survey responses were included if the child attended the target grade and if the ADHD-RS-IV was completed. In case of duplicate responses for the same child across phases, the survey response used to select the child for further studies was selected if applicable. Otherwise, the more complete response was selected, and if both responses were complete, the more recent response was selected. Parents of 465 children in phase I and of 1690 children in phase II responded of which 18 and 51 parents did not complete the ADHD-RS-IV. After adjusting for duplicates between phases (n=42), the final sample consisted of 2044 parent reports of children aged 6–11 years (92 % aged 7–9).

### Background information in the general population sample

Background information regarding grade repetition, special educational need support, referrals, and psychiatric diagnoses was collected. To examine the sample’s representativeness, information on socio-economic status (SES), income, and immigration background was also collected. SES was defined as; (a) the length of education in years on average for both parents/caregivers (range: 0–17.5) and (b) the highest educational level of parents (i.e., if information was available for both parents, the highest education level of the two was reported). Moreover, the annual pre-tax household income was reported and categorized as either (i) below or (ii) equal to or above the average annual pre-tax income per household for families with children in Aarhus of 805,131 DKK (≈107,762 euros) in 2016 [45] operationalized as (i) ≤799,999 or (ii) ≥800,000 DKK. Information on immigration background was based on three questions regarding country of birth adapted from the Danish Health Behaviour in School-aged Children survey (e.g., [46, 47]). The immigration background of the child was categorized as immigrant, descendant (i.e., second generation immigrant), or Danish origin using the definitions employed by Statistics Denmark [48] operationalized and described elsewhere [46, 47].

### Clinical ADHD sample

A clinical sample of children and adolescents with ADHD from a previous study [40] was included to investigate whether the results from the general population could be replicated in a well-diagnosed clinical sample. This sample consisted of 209 children with ADHD referred to the child and adolescent psychiatric hospital in Aarhus from January 2011 to January 2013. For detailed procedures and sample characteristics consult Virring et al. [40]. The ADHD-RS-IV was collected as part of the standard diagnostic assessment at the psychiatric hospital. Since the current study was based on parent report alone, 19 out of 209 children were excluded due to teacher report on the ADHD-RS-IV only (n=6) or unknown respondents (n=13). In addition, 25 children were excluded due to completely or partially (>30 %) missing ADHD-RS-IV. Thus, the ADHD sample included in the current study consisted of 165 children and adolescents aged 6–14 at the time of assessment. Eighty percent had a combined ADHD presentation, 18 % the inattentive presentation, and 2 % the hyperactive/impulsive presentation [40]. All diagnoses were assessed according to DSM-IV criteria using the Development and Well-Being Assessment [49].

### Measures

The ADHD-RS-IV Home version was used in both samples. It has 18 items covering the symptom criteria for ADHD as outlined in DSM-IV [41, 42]. Parents rated the frequency of each symptom within the past six months on a four-point Likert scale ranging from “Never or rarely” (0), “Sometimes” (1), “Often” (2) to “Very often” (3) with higher scores indicating greater degree of ADHD symptomatology and severity [42]. According to the scale developers, nine items cover deficits in attention and can be summed to generate an Inattention subscale (range: 0–27), whereas the remaining nine items cover hyperactivity and impulsivity and can be summed to yield a Hyperactivity/impulsivity subscale (range: 0–27) [42]. A total score can be derived by summing all 18 items (range: 0–54).

### Statistical analyses

#### Factor structure

Confirmatory Factor Analyses (CFAs) were used to investigate the factor structure of ADHD in the general population sample (N=2044) and in the ADHD sample (N=147). First, four correlated models were specified and estimated: a single-factor and unidimensional model including all 18 ADHD-RS-IV items, a two-factor model including an Inattention factor (9 items) and a Hyperactivity/impulsivity factor (9 items) in line with DSM-IV and DSM-5 (Corr-2), and two three-factor models with an Inattention factor (9 items) and separate Hyperactivity (5–6 items) and Impulsivity (3–4 items) factors (two versions of Corr-3). The three-factor models differed solely based on the allocation of the item “talks excessively”. In the DSM-based version of the three-factor model (Corr-3 (talk-hy)), “talks excessively” was included as an indicator of hyperactivity, whereas this item was an indicator of impulsivity in the ICD-10 three-factor model (Corr-3 (talk-imp)). Finally, three orthogonal bifactor alternatives were specified: All with one general ADHD factor, one with two specific factors (Bi-2s) and two with three specific factors differing based on the allocation of the item “talks excessively (Bi-3s (talk-hy) and Bi-3s (talk-imp)).

In the CFAs, items were treated as ordinal and the mean and variance adjusted weighted least squares (WLSMV) estimator was applied in all models. The chi-square statistics (χ^2^) was used to evaluate the global fit of each model with lower and non-significant test statistic indicating adequate fit (p > 0.05). However, because the χ^2^ test is sensitive to sample sizes [50], additional goodness of fit indices were applied and emphasized in the evaluation of model fit, namely the Comparative Fit Index (CFI) [51], the Tucker-Lewis Index (TLI) [52], and the Root Mean Square Error of Approximation (RMSEA) [53] including the 90 % confidence interval. Following the guidelines by Hu and Bentler [54, 55], CFI and TLI values ≥ 0.95 and RMSEA < 0.06 are indicative of good model fit. Fit improvement for nested models (e.g., Corr-2 *vs*. Corr-3 (talk-imp)) was evaluated using the Satorra-Bentler χ^2^ difference test (ΔS-B χ^2^).

Three additional psychometric indices were used to evaluate the bifactor models [16]: the explained common variance (ECV) [56, 57], the omega hierarchical (ω_h_) [58, 59] and subscale (ω_s_) [17, 60] coefficients, and the H coefficient [61, 62]. An ECV value for the general factor approaching or close to 1 (e.g., greater than 0.70) indicate higher degree of unidimensionality and a strong general factor relative to the specific factors [16, 17, 57, 63]. ECV for each specific factor was also reported. The ω_h_ and ω_s_ evaluates model-based reliability (e.g., [16, 60]). Elsewhere, it has been proposed that ω values of at least 0.50 is required as the minimum acceptable level and may even be considered necessary to allow meaningful interpretation of the (specific/general) factor, while ω coefficient values closer to 0.75 are preferred [16, 60, 64]. In addition, ω_h_ values higher than 0.80 indicate that the total score of the rating scale in question can be interpreted as essentially unidimensional because most of the variance are accounted for by a single common source, i.e., by the general factor [16, 63]. The H coefficient evaluates construct replicability and how well a latent (specific/general) factor is captured and represented by a set of items [16, 62, 63]. H values smaller than 0.70 indicate that the latent factor is poorly-defined and likely to be unstable and change across samples and studies, values from 0.70–0.79 are in the acceptable range, whereas values equal to or above 0.80 indicate a good and well-defined latent factor that is likely to show stability across samples and studies [16, 63].

Finally, factor scores were estimated for the best-fitting bifactor model and regression analyses were applied to investigate age and gender differences within each sample.

#### Establishing norms

Norms were estimated based on the summed ADHD-RS-IV scores in the general population sample. The general population had no missing ADHD-RS-IV data, while eighteen children in the ADHD sample had one or two missing items and their scores were therefore calculated as the average of answered and non-missing items multiplied with the number of items. As the distribution of scores was skewed right, percentiles (80, 90, 93, and 98) were presented in line with the US standardization of ADHD-RS-IV [42] for the total and subscale scores, and also for the individual hyperactivity (five items) and impulsivity (four items) scores. Gender-and-age-stratified percentiles were presented along with means, standard deviations, and medians for girls and boys aged 6–9. Differences in ADHD-RS-IV scores, age, and sex distribution between the two samples were analyzed using independent t-tests or chi-square (χ^2^) test.

#### Statistical software

The CFAs were carried out in R version 3.6.1 [65] using the lavaan package [66], and factor scores were derived using the lavPredict function (EBM method). ECV estimates and the ω coefficients (calculated using the model-implied covariance matrix in the denominator) were obtained from the BifactorIndicesCalculator package (omega_H and ECV_SG) [67]. All remaining analyses were conducted in Stata version 16.1 [68].

## RESULTS

### Sample characteristics

Sample characteristics for the general population sample are provided in Table 1. The general population sample consisted of 1099 boys and 945 girls with a mean age of 8.23 (SD=0.92). There was an almost equal distribution of 1st (36.2 %), 2nd (34.9 %), and 3rd (28.9 %) graders. Around 2 % had repeated a grade level and 7.2 % received some sort of special educational need support (i.e., educational support, teaching assistance, or attending a special class/school). The majority were classified as being of Danish origin (95.0 %), but children with descendant (4.3 %) and immigrant background (0.7 %) were also represented. Parents had on average 15.07 (SD=2.01) years of education with medium-cycle and long-cycle higher education as the most common educational level. Five percent of the children had been referred to the child and adolescent psychiatric hospital and 3.8 % were reported to have at least one psychiatric disorder. Almost 2 % were reported to have an ADHD/ADD diagnosis.

**Table 1.**
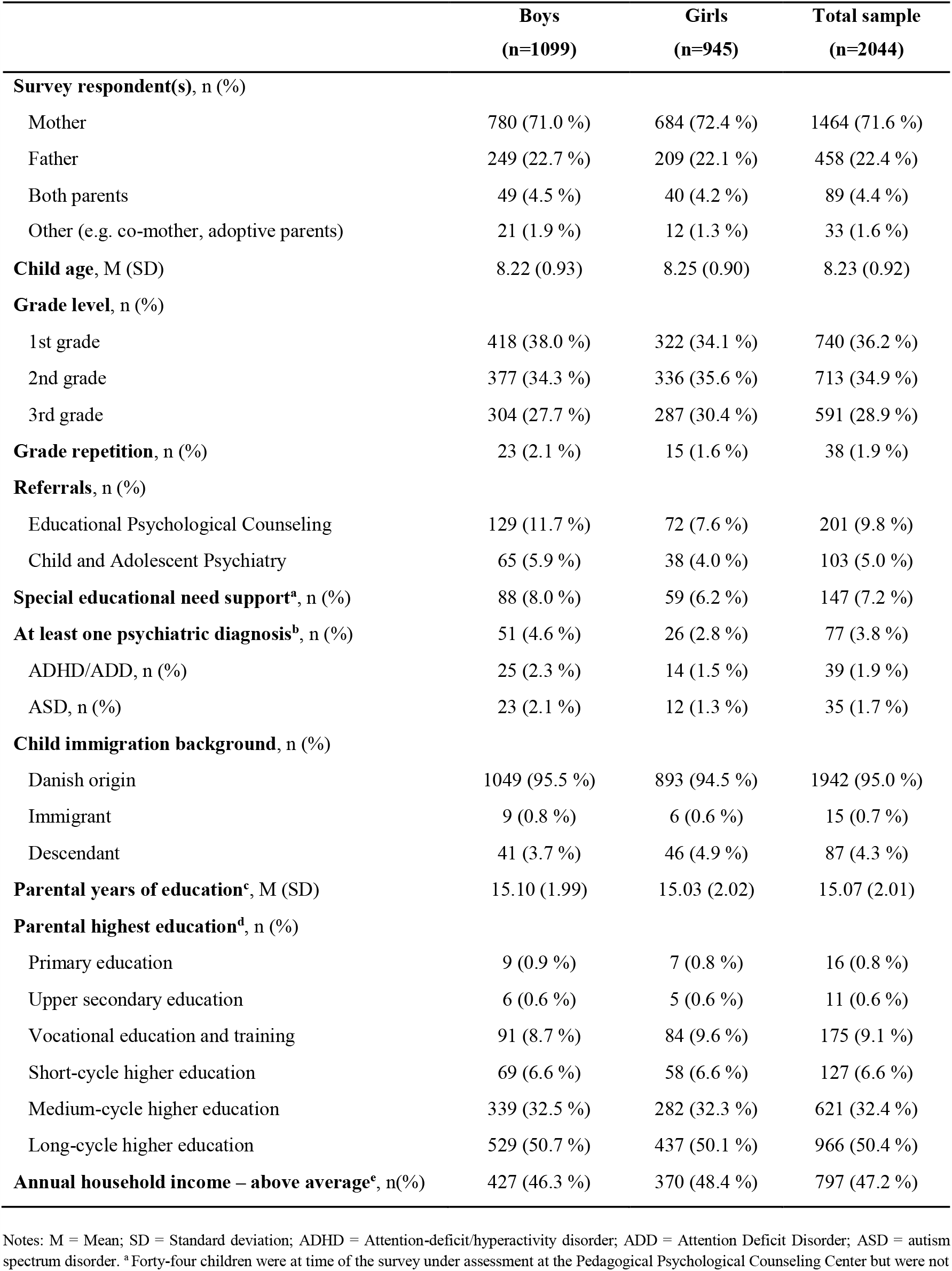
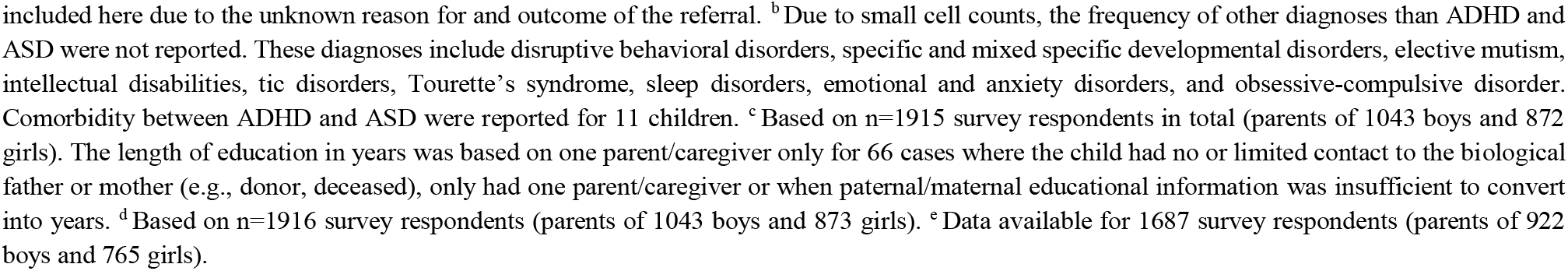
Sample characteristics of the general population sample

|  | **Boys**<br>**(n=1099)** | **Girls**<br>**(n=945)** | **Total sample**<br>**(n=2044)** |
| --- | --- | --- | --- |
| **Survey respondent(s), n (%)** |  |  |  |
| Mother | 780 (71.0 %) | 684 (72.4 %) | 1464 (71.6 %) |
| Father | 249 (22.7 %) | 209 (22.1 %) | 458 (22.4 %) |
| Both parents | 49 (4.5 %) | 40 (4.2 %) | 89 (4.4 %) |
| Other (e.g. co-mother, adoptive parents) | 21 (1.9 %) | 12 (1.3 %) | 33 (1.6 %) |
| **Child age, M (SD)** | 8.22 (0.93) | 8.25 (0.90) | 8.23 (0.92) |
| **Grade level, n (%)** |  |  |  |
| 1st grade | 418 (38.0 %) | 322 (34.1 %) | 740 (36.2 %) |
| 2nd grade | 377 (34.3 %) | 336 (35.6 %) | 713 (34.9 %) |
| 3rd grade | 304 (27.7 %) | 287 (30.4 %) | 591 (28.9 %) |
| **Grade repetition, n (%)** | 23 (2.1 %) | 15 (1.6 %) | 38 (1.9 %) |
| **Referrals, n (%)** |  |  |  |
| Educational Psychological Counseling | 129 (11.7 %) | 72 (7.6 %) | 201 (9.8 %) |
| Child and Adolescent Psychiatry | 65 (5.9 %) | 38 (4.0 %) | 103 (5.0 %) |
| **Special educational need support<sup>a</sup>, n (%)** | 88 (8.0 %) | 59 (6.2 %) | 147 (7.2 %) |
| **At least one psychiatric diagnosis<sup>b</sup>, n (%)** | 51 (4.6 %) | 26 (2.8 %) | 77 (3.8 %) |
| ADHD/ADD, n (%) | 25 (2.3 %) | 14 (1.5 %) | 39 (1.9 %) |
| ASD, n (%) | 23 (2.1 %) | 12 (1.3 %) | 35 (1.7 %) |
| **Child immigration background, n (%)** |  |  |  |
| Danish origin | 1049 (95.5 %) | 893 (94.5 %) | 1942 (95.0 %) |
| Immigrant | 9 (0.8 %) | 6 (0.6 %) | 15 (0.7 %) |
| Descendant | 41 (3.7 %) | 46 (4.9 %) | 87 (4.3 %) |
| **Parental years of education<sup>c</sup>, M (SD)** | 15.10 (1.99) | 15.03 (2.02) | 15.07 (2.01) |
| **Parental highest education<sup>d</sup>, n (%)** |  |  |  |
| Primary education | 9 (0.9 %) | 7 (0.8 %) | 16 (0.8 %) |
| Upper secondary education | 6 (0.6 %) | 5 (0.6 %) | 11 (0.6 %) |
| Vocational education and training | 91 (8.7 %) | 84 (9.6 %) | 175 (9.1 %) |
| Short-cycle higher education | 69 (6.6 %) | 58 (6.6 %) | 127 (6.6 %) |
| Medium-cycle higher education | 339 (32.5 %) | 282 (32.3 %) | 621 (32.4 %) |
| Long-cycle higher education | 529 (50.7 %) | 437 (50.1 %) | 966 (50.4 %) |
| **Annual household income – above average<sup>e</sup>, n(%)** | 427 (46.3 %) | 370 (48.4 %) | 797 (47.2 %) |

Notes: M = Mean; SD = Standard deviation; ADHD = Attention-deficit/hyperactivity disorder; ADD = Attention Deficit Disorder; ASD = autism spectrum disorder. <sup>a</sup>Forty-four children were at time of the survey under assessment at the Pedagogical Psychological Counseling Center but were not
included here due to the unknown reason for and outcome of the referral. <sup>b</sup> Due to small cell counts, the frequency of other diagnoses than ADHD and ASD were not reported. These diagnoses include disruptive behavioral disorders, specific and mixed specific developmental disorders, elective mutism, intellectual disabilities, tic disorders, Tourette's syndrome, sleep disorders, emotional and anxiety disorders, and obsessive-compulsive disorder. Comorbidity between ADHD and ASD were reported for 11 children. <sup>c</sup> Based on n=1915 survey respondents in total (parents of 1043 boys and 872 girls). The length of education in years was based on one parent/caregiver only for 66 cases where the child had no or limited contact to the biological father or mother (e.g., donor, deceased), only had one parent/caregiver or when paternal/maternal educational information was insufficient to convert into years. <sup>d</sup> Based on n=1916 survey respondents (parents of 1043 boys and 873 girls). <sup>e</sup> Data available for 1687 survey respondents (parents of 922 boys and 765 girls).

The clinical ADHD sample consisted of 129 boys and 36 girls with a mean age of 9.76 (SD=1.88). In this sample, 53.3 % had at least one comorbid disorder (primary comorbidity: 59.1 % externalizing disorder, 23.9 % internalizing disorder, 6.8 % autism spectrum disorder, and 10.2 % tics disorder / Tourette’s syndrome), and 14.5 % had more than one comorbid disorder. The ADHD sample was on average 1.53 (95 % CI: 1.23–1.82) years older (*t*(170.31)=10.30, *p*<0.0001) and included a higher proportion of boys (78.2 % *vs*. 53.8 %; *χ*^2^(1)=36.86, *p*<0.001) than the general population sample.

### Factor structure

Fit indices for all CFA models are presented in Table 2 and bifactor indices are shown in Table 3. Standardized factor loadings for all models are available in Online Resource 2.

**Table 2.** Fit indices for the CFAs

| Model | $\chi^2$ (df) | CFI | TLI | RMSEA [90 % CI] |
| --- | --- | --- | --- | --- |
| **General population sample (N=2044)** |  |  |  |  |
| Single-factor | 3040.66 (135)** | 0.976 | 0.973 | 0.103 [0.099-0.106] |
| Corr-2: IN, HY/IMP <sup>a</sup> | 1356.92 (134)** | 0.990 | 0.989 | 0.067 [0.064-0.070] |
| Corr-3 (talk-hy): IN, HY, IMP <sup>b</sup> | 1054.29 (132)** | 0.992 | 0.991 | 0.058 [0.055-0.062] |
| Corr-3 (talk-imp): IN, HY, IMP <sup>c</sup> | 737.67 (132)** | 0.995 | 0.994 | 0.047 [0.044-0.051] |
| Bi-2s: G, IN<sub>s</sub>, HY/IMP<sub>s</sub> | 755.47 (117)** | 0.995 | 0.993 | 0.052 [0.048-0.055] |
| Bi-3s (talk-hy): G, IN<sub>s</sub>, HY<sub>s</sub>, IMP<sub>s</sub> | 806.14 (117)** | 0.970 | 0.961 | 0.054 [0.050-0.057] |
| Bi-3s (talk-imp): G, IN<sub>s</sub>, HY<sub>s</sub>, IMP<sub>s</sub> | 586.19 (117)** | 0.996 | 0.995 | 0.044 [0.041-0.048] |
| **ADHD sample (N=147)** |  |  |  |  |
| Single-factor | 353.75 (135)** | 0.958 | 0.952 | 0.105 [0.092-0.119] |
| Corr-2: IN, HY/IMP <sup>d</sup> | 198.35 (134)** | 0.988 | 0.986 | 0.057 [0.040-0.074] |
| Corr-3 (talk-hy): IN, HY, IMP <sup>e</sup> | 181.55 (132)* | 0.990 | 0.989 | 0.051 [0.031-0.068] |
| Corr-3 (talk-imp): IN, HY, IMP <sup>f</sup> | 156.71 (132) | 0.995 | 0.994 | 0.036 [0.000-0.056] |
| Bi-2s: G, IN<sub>s</sub>, HY/IMP<sub>s</sub> | 101.27 (117) | 1.000 | 1.000 | 0.000 [0.000-0.025] |
| Bi-3s (talk-hy): G, IN<sub>s</sub>, HY<sub>s</sub>, IMP<sub>s</sub> | 108.58 (117) | 1.000 | 1.000 | 0.000 [0.000-0.034] |
| Bi-3s (talk-imp): G, IN<sub>s</sub>, HY<sub>s</sub>, IMP<sub>s</sub> | 101.79 (117) | 1.000 | 1.000 | 0.000 [0.000-0.025] |

Notes: ADHD = Attention-deficit/hyperactivity disorder; CFA = Confirmatory Factor Analysis; df = degree of freedom; CFI = Comparative Fit Index; TLI = Tucker–Lewis Index; RMSEA = root mean square error of approximation; CI = confidence interval; Corr-2 = Correlated two-factor model; Corr-3 = Correlated three-factor model; talk-hy = The item “talks excessively” is specified to load on the Hyperactivity factor; talk-imp = The item “talks excessively” is specified to load on the Impulsivity factor; Bi-2s = Bifactor model with one general and two specific factors; Bi-3s = Bifactor model with one general and three specific factors; G = General ADHD factor; IN<sub>(s)</sub> = (Specific) Inattention factor; HY-IMP<sub>(s)</sub> = (Specific) Hyperactivity/impulsivity factor; HY<sub>(s)</sub> = (Specific) Hyperactivity factor; IMP<sub>(s)</sub> = (Specific) Impulsivity factor. <sup>a</sup> The correlation between IN and HY-IMP=0.78. <sup>b</sup> The correlations between factors were: IN-HY=0.80, IN-IMP=0.68, and HY-IMP=0.85. <sup>c</sup> The correlations between factors were: IN-HY=0.80, IN-IMP=0.67, and HY-IMP=0.78. <sup>d</sup> Correlation between IN and HY-IMP=0.68. <sup>e</sup> The correlation between factors were: IN-HY=0.64; IN-IMP=0.68, and HY-IMP=0.85. <sup>f</sup> The correlation between factors were: IN-HY=0.63; IN-IMP=0.67, and HY-IMP=0.76. \*\*p<0.001. \*p<0.01.

**Table 3.**
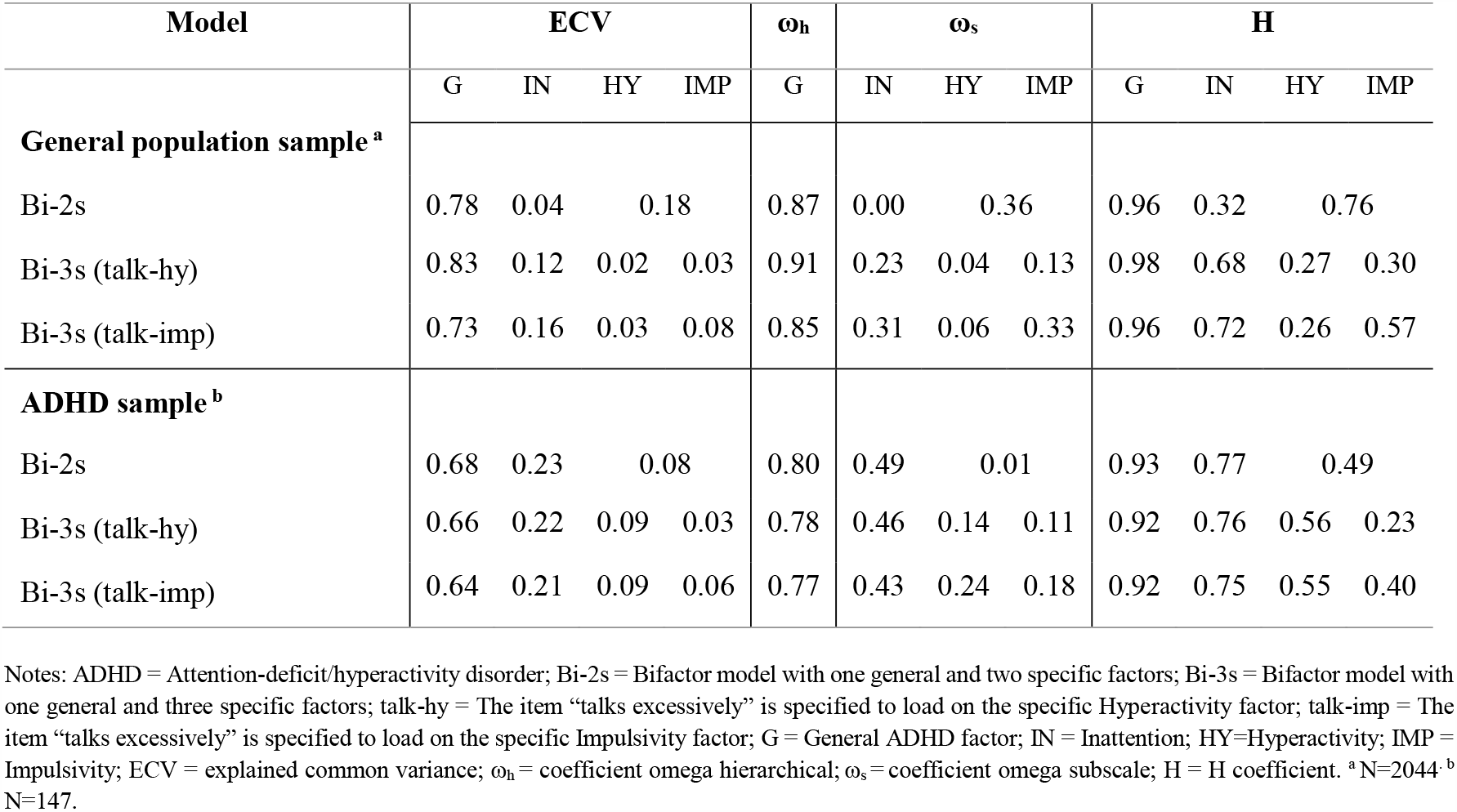
Bifactor indices

#### General population sample

All correlated factor models showed good model fit in terms of CFI and TLI (all >0.95, see Table 2). Only the correlated three-factor models fit the data well according to RMSEA (<0.06). Difference testing also favored the correlated three-factor models over the two-factor model (Corr-2 *vs*. Corr-3 (talk-hy): ΔS-B χ^2^(2)=165.36, *p*<0.0001; Corr-2 *vs*. Corr-3 (talk-imp): ΔS-B χ^2^(2)=265.54, *p*<0.0001), although the two-factor model fit better than the single-factor model (ΔS-B χ^2^(1)=429.20, *p*<0.0001). The latent factors of the models were highly correlated (ranging from 0.67 to 0.85, revisit Table 2).

All bifactor models fit the data well (all CFI/TLI > 0.95 and RMSEA < 0.06; for non-significant and/or negative factor loadings see Online Resource 2). The bifactor indices suggested that all bifactor models had a strong and reliable general factor and relatively weak specific factors (see Table 3). The general factor explained between 73%–83% of the common variance (Bi-2s and Bi-3s models), while the specific Inattention factor explained 4–16 % (Bi-2s and both Bi-3s), the Hyperactivity/impulsivity factor explained 18% (Bi-2s), and the separate Hyperactivity and Impulsivity factors explained 2–3 % and 3–8 % of the common variance (Bi-3s models), respectively. Only the general factors reached an ω coefficient value above the threshold allowing for meaningful interpretation and this coefficient even indicated unidimensionality (ω_h_>0.80). Finally, H values were in the good range for the general factor (H=0.96–0.98), in the acceptable range for the specific Inattention factor (H=0.72) in the Bi-3s (talk-imp) model only (but not in Bi-2s and Bi-3s (talk-hy)), and below the acceptable range for the specific Impulsivity (H=0.30–0.57) and especially for the Hyperactivity factor (H=0.26–0.27) in the Bi-3s models. The specific Hyperactivity/impulsivity factor in the Bi-2s model was within the acceptable range (H=0.76).

Based on fit indices alone, the correlated ICD-10 three-factor, Corr-3 (talk-imp), and the bifactor version of this model, Bi-3s (talk-imp), were marginally better than the remaining models. Difference testing clearly supported a three-factor model as the better model (compared to a two-factor version). However, the strong interrelatedness between factors and results from the bifactor indices suggested that a strong general factor accounted for most of the common variance in ADHD-RS-IV items with two or three (talk-imp) weak specific factors. Regarding “talks excessively”, this item had a significant factor loading (λ=0.50) on the Impulsivity factor in the Bi-3s (talk-imp) model, but a very low, negative, and non-significant (λ=-0.07) factor loading on the Hyperactivity factor in the Bi-3s (talk-hy) model.

#### Clinical ADHD sample

With the exception of the single-factor model, all correlated factor models provided a good fit to the data (see Table 2). Difference testing suggested that the correlated two-factor model was a significant improvement in fit over the single-factor model (ΔS-B χ^2^(1)=33.63, *p*<0.0001), but that the three-factor models were a significant improvement compared to the two-factor model (Corr-2 *vs*. Corr-3 (talk-hy): ΔS-B χ^2^(2)=9.49, *p*<0.01; Corr-2 *vs*. Corr-3 (talk-imp): ΔS-B χ^2^(2)=17.71, *p*<0.001). In addition, only the Corr-3 (talk-imp) model had a non-significant χ^2^ statistics. Again, the latent factors of the two- and three-factor models were highly correlated (ranging from 0.63 to 0.85).

All bifactor models in the ADHD sample fit the data well (see Table 2; see Online Resource 2 for non-significant and/or negative factor loadings). The bifactor indices suggested the presence of a relatively strong general factor in all models explaining between 64%–68% of the common variance and specific factors each explaining only 3–23% (revisit Table 3). The general factor demonstrated a good level of reliability (ω_h_=0.77–0.80) and good construct replicability (all H>0.80) in all models. The specific Inattention factor in the ADHD sample also had acceptable construct replicability (H=0.75–0.77) and approached acceptable model-based reliability (ω_s_=0.43–0.49). Specific factors involving hyperactivity and impulsivity (separately or in combination) had psychometric bifactor indices below recommended thresholds indicating weak specific factor(s).

Based on fit indices alone, the correlated ICD-based three-factor, Corr-3 (talk-imp), and all the bifactor models were marginally better than the remaining models. Difference testing supported a three-factor solution compared to a two-factor version. Again, factors in the correlated models were highly correlated and results from the additional bifactor indices suggested that a strong general factor accounted for most of the common variance in items, although some support for a specific Inattention factor was also found in this sample. In this sample, the item “talks excessively” had a significant loading (λ=0.46) on the Impulsivity factor in the Bi-3s (talk-imp) model, but a negative and non-significant loading on the Hyperactivity factor (λ=-0.23) in the Bi-3s (talk-hy) model.

### Gender and age differences in factor scores

Factor scores from the Bi-3s (talk-imp) model were used to examine age and gender differences. In the general population sample, boys had higher factor scores on the general factor and two specific factors (*F*_*general*_(1,2042)=63.76, *p*<0.0001; *F*_*Inattention*_(1,2042)=10.41, *p*=0.001; *F*_*Hyperactivity*_(1,2042)=9.23, *p*=0.002) compared to girls. Girls were found to have higher Impulsivity factor scores compared to boys (*F*(1,2042)=33.24, *p*<0.0001). On average, older children had higher Inattention (*F*(1,2042)=16.40, *p*<0.001) and lower Hyperactivity factor scores (*F*(1,2042)=20.57, *p*<0.0001). There was no significant age effect on the general (*F*(1,2042)=2.93, *p*=0.087) or the Impulsivity factor score (*F*(1,2042)=2.07, *p*=0.150). In the ADHD sample, there were no gender differences on the general (*F*(1,145)=0.02, *p*=0.895), Inattention (*F*(1,145)=2.63, *p*=0.107), or Impulsivity factor scores (*F*(1,145)=1.73, *p*=0.190), while boys had higher Hyperactivity factor scores compared to girls (*F*(1,145)=12.14, *p*<0.001). On average, older children had greater Inattention (*F*(1,145)=8.66, *p*=0.004) and lower general factor scores (*F*(1,145)=5.90, *p*=0.016). There was no significant age effect on the Hyperactivity (*F*(1,145)=2.38, *p*=0.125) or Impulsivity factor score (*F*(1,145)=0.19, *p*=0.665). Moreover, no significant interaction effects of age and gender on the factor scores emerged in any of the samples (analyses not shown).

### Normative data based on summed scores

Table 4 presents gender-and-age-stratified normative data for the general population sample (age 6–9) based on summed ADHD-RS-IV scores. Means, standard deviations, and medians for the ADHD-RS-IV scores stratified according to gender and age in the ADHD sample and comparison on summed ADHD-RS-IV scores between samples are available in Online Resource 3. In short, children in the ADHD sample were reported to have significantly higher scores on all summed ADHD-RS-IV scores compared to the general population sample.

**Table 4.** Normative ADHD-RS-IV Home version data for schoolchildren aged 6–9 from the general population

|  | n | M | SD | Mdn | 80th<br>percentile | 90th<br>percentile | 93th<br>percentile | 98th<br>percentile |
| --- | --- | --- | --- | --- | --- | --- | --- | --- |
| **Girls aged 6–9** |  |  |  |  |  |  |  |  |
| Total score | 879 | 8.70 | 7.95 | 7 | 13 | 18 | 21 | 33 |
| Inattention | 879 | 4.44 | 4.62 | 3 | 7 | 10 | 12 | 19 |
| Hyperactivity/impulsivity | 879 | 4.26 | 4.07 | 3 | 7 | 9 | 11 | 17 |
| Hyperactivity alone<sup>a</sup> | 879 | 1.75 | 2.28 | 1 | 3 | 5 | 5 | 9 |
| Impulsivity alone<sup>b</sup> | 879 | 2.51 | 2.27 | 2 | 4 | 5 | 6 | 9 |
| **Boys aged 6–9** |  |  |  |  |  |  |  |  |
| Total score | 1016 | 11.11 | 8.79 | 9 | 17 | 23 | 26 | 36 |
| Inattention | 1016 | 5.91 | 4.95 | 5 | 10 | 13 | 14 | 20 |
| Hyperactivity/impulsivity | 1016 | 5.20 | 4.62 | 4 | 8 | 12 | 13 | 19 |
| Hyperactivity alone<sup>a</sup> | 1016 | 2.59 | 2.75 | 2 | 4 | 6 | 8 | 11 |
| Impulsivity alone<sup>b</sup> | 1016 | 2.62 | 2.35 | 2 | 4 | 6 | 6 | 9 |

Notes: n = sample size; M = Mean; SD = Standard deviation; Mdn = Median; ADHD-RS-IV = Attention-Deficit/Hyperactivity Disorder–Rating Scale IV. <sup>a</sup> This score is comprised by five items (i.e., “fidgets/squirms”, “leaves seat”, “runs about”, “difficulty playing quietly”, and “on the go”). <sup>b</sup> This score is comprised by four items (i.e., “talks excessively”, “blurts out answers”, “difficulty awaiting turns”, and “interrupts or intrudes”).

## DISCUSSION

This study examined the underlying structure of ADHD in a large general population sample of schoolchildren and in a clinical sample of children with ADHD. While all models except for the single-factor model (both samples) and the correlated two-factor model (general population sample) provided good fit, the correlated three-factor model with “talks excessively” included in the Impulsivity factor and the bifactor version of this model were found to be marginally better in the general population sample. Results were largely replicated in the ADHD sample, although the fit indices indicated excellent fit for all bifactor models, possibly due to the smaller sample size in combination with the high number of parameters in these models.

### Bifactor *vs*. correlated models

In keeping with much prior research (e.g., [18–26]), the traditional goodness of fit indices showed that the bifactor models provided a better fit than the correlated models in both samples. Recently, some concerns have been raised about relying on or overemphasizing traditional goodness of fit indices as these indices may favor the less constrained model possessing more parameters such as the bifactor model and due to the potential tendency to overfit data ascribed to the bifactor models [37–39]. Instead, reliance on bifactor-specific psychometric indices for model evaluation has been recommended [16, 38]. When we applied such indices in the current study, a strong and well-defined general ADHD factor emerged. The specific factors, on the other hand, did not appear to contribute much to the model above and beyond the general ADHD factor. In fact, these factors may represent residual error or nuisance variance with questionable clinical utility and practical value [15, 26]. Specifically, the combined or separate specific Hyperactivity and Impulsivity factor(s) generally proved to be weak and poorly-defined in both samples. The specific Inattention factor contributed with more unique information and was more reliable in the clinical sample than in the general population sample. That is, the bifactor indices for this factor were above or approached acceptable thresholds in the ADHD sample, but only reached acceptable construct replicability in one of the bifactor models (Bi-3s (talk-imp)) in the general population sample but otherwise presented as a weak factor with all inattention items having a higher loading on the general than on the specific Inattention factor. These results are in line with a review that estimated bifactor indices for 31 bifactor models of ADHD derived from 18 studies [16]. They also found evidence for a strong general factor with psychometrical meaningful and reliable properties while the specific factors were shown to be weak and poorly-defined (especially Hyperactivity/impulsivity), although the specific Inattention factor appeared to have some merit in clinical (but not in general population) samples [16].

While the bifactor models were characterized by relatively weak specific factors [16], factors in all correlated models were found to be highly correlated (*r*=0.63–0.85). This is in line with previous studies reporting the association between the Hyperactivity and Impulsivity factors to be higher than 0.80 in the three-factor model [4–6, 15, 20–22, 27, 30, 69], and ranging from 0.56 to 0.98 for the Inattention and the Hyperactivity/impulsivity factors in the two-factor model [6, 9, 20, 21, 27, 30, 31, 69]. This degree of interrelatedness questions whether the factors should be separated and may blur or even hamper a clear-cut interpretation of the correlated model [16, 22]. As the bifactor indices showed that most of the variance in each individual ADHD-RS-IV item is shared with the other ADHD-RS-IV items consistent with previous studies (e.g. [16, 26]), a bifactor model with a strong general factor and two or three weaker specific factors appears to be a better representation of the factor structure of ADHD irrespective of the potentially inflated goodness of fit indices due to the model complexity.

### Talks excessively

To date, studies have varied with regard to the specification of the symptom “talks excessively” when examining the structure of ADHD – sometimes including it as a symptom of hyperactivity, sometimes of impulsivity. Three studies of adults from a community [25], a psychiatric outpatients [33] and an ADHD sample [24] have previously investigated competing three-(bi)factor models differing on the allocation of “talks excessively”. These studies found support for the ICD-10 organization of hyperactivity and impulsivity symptoms. Here we extend these findings to young schoolchildren in the general population and to children and adolescents with ADHD. Of the investigated correlated models, the three-factor model where “talks excessively” was included as an impulsivity item was supported as a marginally better model in both samples. The same result was obtained for the bifactor version of this model in the general population sample, but a similar fit of the bifactor models was found in the ADHD sample for reasons stated above. The bifactor loadings of this item in both samples with positive and significant loadings on the specific Impulsivity factor compared to non-significant and negative loadings on the specific Hyperactivity factor further support the “talks excessively” item as belonging to the impulsivity construct.

### Two or three (specific) factors?

Our results generally (albeit marginally) supported three rather than two factors, and the ICD-10 organization of these factors. We speculated whether the inconsistent findings from previous studies regarding the number of specific factors (e.g., two *vs*. three) may be explained by the differences in how these factors have been specified. Indeed, studies comparing the Bi-2s and Bi-3s (talk-imp) models have supported the latter [24, 25, 32, 33] or an incomplete bifactor model with specific factors for Inattention and Impulsivity only [23, 36], whereas more mixed findings have been reported when compared to the Bi-3s (talk-hyp) model. These studies, including studies not directly reporting which factor the item “talks excessively” belonged to but that focused on or used a DSM-IV/5 specific diagnostic interview or rating scale, have supported the (i) Bi-2s model [18, 19, 70–72], the (ii) Bi-3s (talk-hyp) model [21], or (iii) both models [34]. Additionally, two studies proposed a (iv) second-order model [6] or a (v) correlated three-factor model [35] as the final model instead of the bifactor models. In the present study, the Bi-3s (talk-hyp) model generally had a lower fit than all models in the general population sample or showed equivalent degree of good fit as the remaining bifactor models in the ADHD sample. Thus, the results would have been less clear if only the Bi-3s (talk-hyp) model had been tested, and future studies should include models based on the ICD-10 structure.

### Clinical implications

Although the bifactor models with a strong general factor may appear more in line with the unitary construct of ADHD previously applied in DSM-III-R [26] than with the two or three separate symptom dimensions currently applied in DSM-5 and ICD-10, a unidimensional single-factor model was not supported in the present study. The fact that the general and the specific factor scores were found to display different patterns of association with gender and age further supports the conclusion that the underlying structure is not simply unidimensional. This is in line with recent suggestions that at least one specific factor in addition to inattention is needed to model the residual dependence between items [15]. However, and as suggested elsewhere [23, 36], the bifactor model seems to support a more dimensional diagnostic approach and an emphasis on the overall level of ADHD rather than counting and adding up symptoms within each of the two or three interrelated symptom dimensions.

Based on the findings from the bifactor studies, the use of the summed total score has been recommended [16, 20, 31, 36] as well as the summed Inattention score in clinical samples [16]. However, summed scores, such as the ADHD-RS-IV total score and especially the subscale scores, are unweighted and not synonymous with the orthogonal general or the specific factor(s) in a bifactor model, whereas weighted factor scores are [26, 69, 72]. Nonetheless, Willoughby et al. [31] found that a general factor score derived from a bifactor model in 1st graders was strongly associated with both the summed total, inattention, and hyperactivity/impulsivity scores and with symptom counts using the Disruptive Behavior Disorders Rating Scale [73]. The specific Inattention and Hyperactivity/impulsivity factor scores, in turn, only explained modest variation in summed scores [31]. Although we did provide norms for the ADHD-RS-IV summed subscale scores proposed by the scale developers, we suggest attaching more importance to the ADHD-RS-IV total score in clinical practice to approximate the general factor indicative of an overall liability for ADHD [26]. In addition to using the total score or alternatively an overall symptom count, Goh et al. [72] proposed the implementation of specific factor scores in clinical practice. Future research should investigate the clinical usefulness and relevance of bifactor scores [69], and whether weighted bifactor scores could be used to establish norms enabling the clinical assessment of a child’s placement on the general and specific (Inattention) factor(s). If so, then computerized algorithms are needed [16, 72] to ease the practical and clinical interpretation of the factor score profiles once investigated throughout.

For now, only normative data for summed ADHD-RS-IV scores was presented in the present study. The scores defining the percentiles (and T-score if calculated) were generally found to be higher compared to the non-published percentiles made available by the authors of the Danish validation study of ADHD-RS-IV [43]. This may be due to the considerable larger sample size in the current study (e.g., 1016 in the present study *vs*. 104 boys aged 6–9 in the validation study [43]) that may better capture the dispersion of ADHD-RS-IV scores in the population. Overall, our results indicate that the Danish norm currently applied in clinical practice may result in over-identification of children aged 6–9 as having clinically significant ADHD symptoms.

### Strengths and limitations

The strengths of the study are the large general population sample (N=2044) with a narrow age band, the use of a validated ADHD questionnaire, and the inclusion of a well-diagnosed ADHD sample to allow for comparison, although the sample size (N=147–165) was relatively small.

Information on the (i) highest completed education, (ii) income, and (iii) immigration status were collected in the general population sample to compare with statistics available at Statistics Denmark. The distribution of the (i) highest completed education of individuals aged 30–49 in Aarhus by October 1st 2016 was: 12.7 % primary education, 6.8 % upper secondary education or qualifying educational programs, 23.7 % vocational education & training, 6.7 % short-cycle, 23.6 % medium-cycle, and 26.5 % long-cycle higher education (or PhD) [74]. In the general population sample, the highest completed education (based on the highest level of one out of two parents) was skewed towards medium- and long-cycle higher education. However, if available information for all parents were included (n=3776), the distribution, in the same order as above, was 3.3 %, 2.0 %, 15.8 %, 9.2 %, 33.2 %, and 36.3 % (and 0.1 % no education), thereby representing the various education levels and slightly approaching the distribution in Aarhus, although fewer with primary, upper secondary, and vocational education were represented. Regarding (ii) income, approximately half of the survey responders had a pre-tax household income below and the other half above the average income of families with children in Aarhus [45]. Finally, (iii) the ancestry of children aged 7–9 by the second quarter of 2016 in Aarhus were 83.1 % with Danish origin, 13.7 % descendants, and 3.2 % immigrants according to Statistics Denmark [44]. Although children with descendant and immigrant background were represented, they only comprised 5 % in the general population sample. This may partially be due to the limited Danish language proficiency of some parents of children with descendant/immigrant background. Moreover, in the absence of information regarding citizenship, Danish origin was defined as having at least one parent born in Denmark irrespective of the birth country of the child [46, 47], potentially including some children with descendant status as Danish origin. Overall, the sample seem representative, although comprising fewer children with descendants/immigrant background and parents with lower SES.

## Conclusion

Overall, our study supported a bifactor model with a general factor and two or three (ICD-10) specific factors. Scrutiny of the bifactor-specific ancillary indices suggested the presence of a strong general factor but weak specific factors, although there was some support for a specific Inattention factor in the ADHD sample. We recommend applying the ADHD-RS-IV total score in clinical practice until more research on the clinical usefulness of factor scores have been conducted.

### Ethical standards

The survey was registered with and approved by the Danish Data Protection Agency (1-16-02-542-15), and the data was processed and stored in accordance with the European Union General Data Protection Regulation. Ethical review board approval is not required for survey-based studies in Denmark. The study by Virring et al. [40] providing data for this analysis was approved by the Danish Regional Ethics Committee (M-20100231) and the Danish Data Protection Agency (2010-41-5472).

## Data Availability

The participants have not agreed to the data being shared. For this reason, the data is only available to the authors of this manuscript.

## Acknowledgement

The authors thank the Department for Children and Young People in Aarhus Municipality, Denmark, for announcing the survey in their newsletter to school leaders, the participating schools and school leaders who helped disseminate information about and distributing the link to the electronic survey, and all parents who participated in the survey.

## Funding

The project was funded by the Novo Nordisk Foundation (NNF15OC0017706); the Lundbeck Foundation (R208-2015-3329); the A.P. Møller Foundation for the Advancement of Medical Science (15-15); the Research Foundation for the Center for the Child and Adolescent Psychiatry, Central Denmark Region (BUC16-TWA, BUC17-TWA); the Psychiatric Research Foundation for the Central Denmark Region (no grant number); the Riisfort Fonden (no grant number); and the Brødrene Hartmanns Fond (R69-A28284-B23205). The funders had no involvement in the design of the study; the data collection; the statistical analyses and interpretation of the results; or the writing and approval of and the decision to submit the manuscript for publication.

## Conflicts of interest

Per Hove Thomsen has received speaker’s fee from Medice and Shire within the last three years. Edmund J.S. Sonuga-Barke has received speaker’s fee from Shire and served as consultant to Neurotech Solutions within the last three years. Trine Wigh Arildskov, Anne Virring, Rikke Lambek, Anders Helles Carlsen, and Søren D. Østergaard declare that they have no conflicts of interest.

## Online Resource 1: Sample flowchart

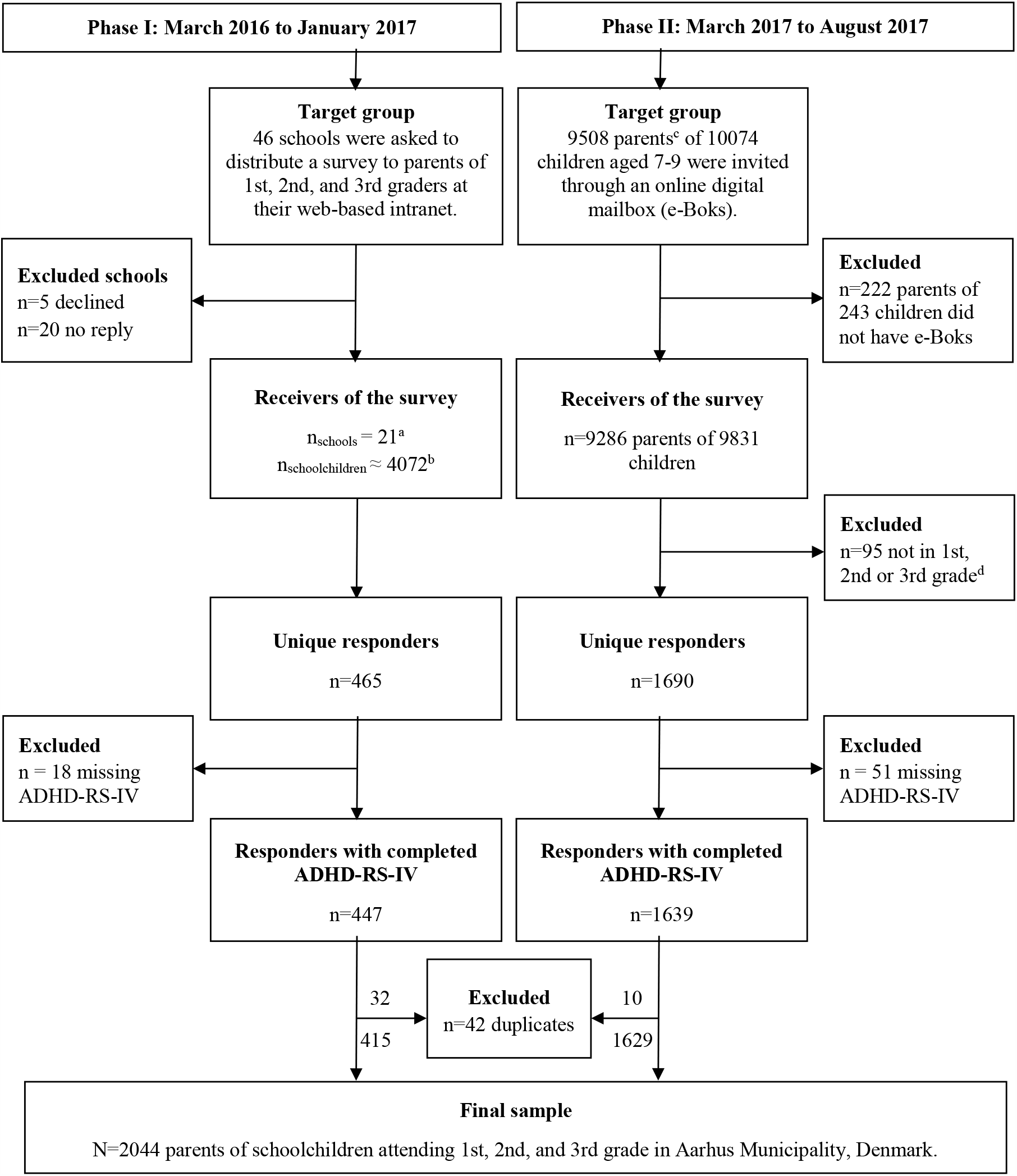
Notes: ^a^ The invitation to the survey was posted at the web-based intranet facility at 20 schools and handed out as a paper version on one school. ^b^ Estimated based on (a) the pupil numbers informed by the school leader, (b) the pupil number available at the webpage of the schools, or (c) based on the pupil number from the previous school year when the children were attending kindergarten, 1st or 2nd grade derived from the quality reports available at the school webpage. ^c^ If the child had more than one parent at the same address, one parent was randomly selected to receive the survey. ^d^ The survey ended immediately if the parents had stated the grade level of the child as “other” than 1^st^, 2^nd^ and 3^rd^ grade.

## Online Resource 2: Standardized factor loadings

**Figure S1.**
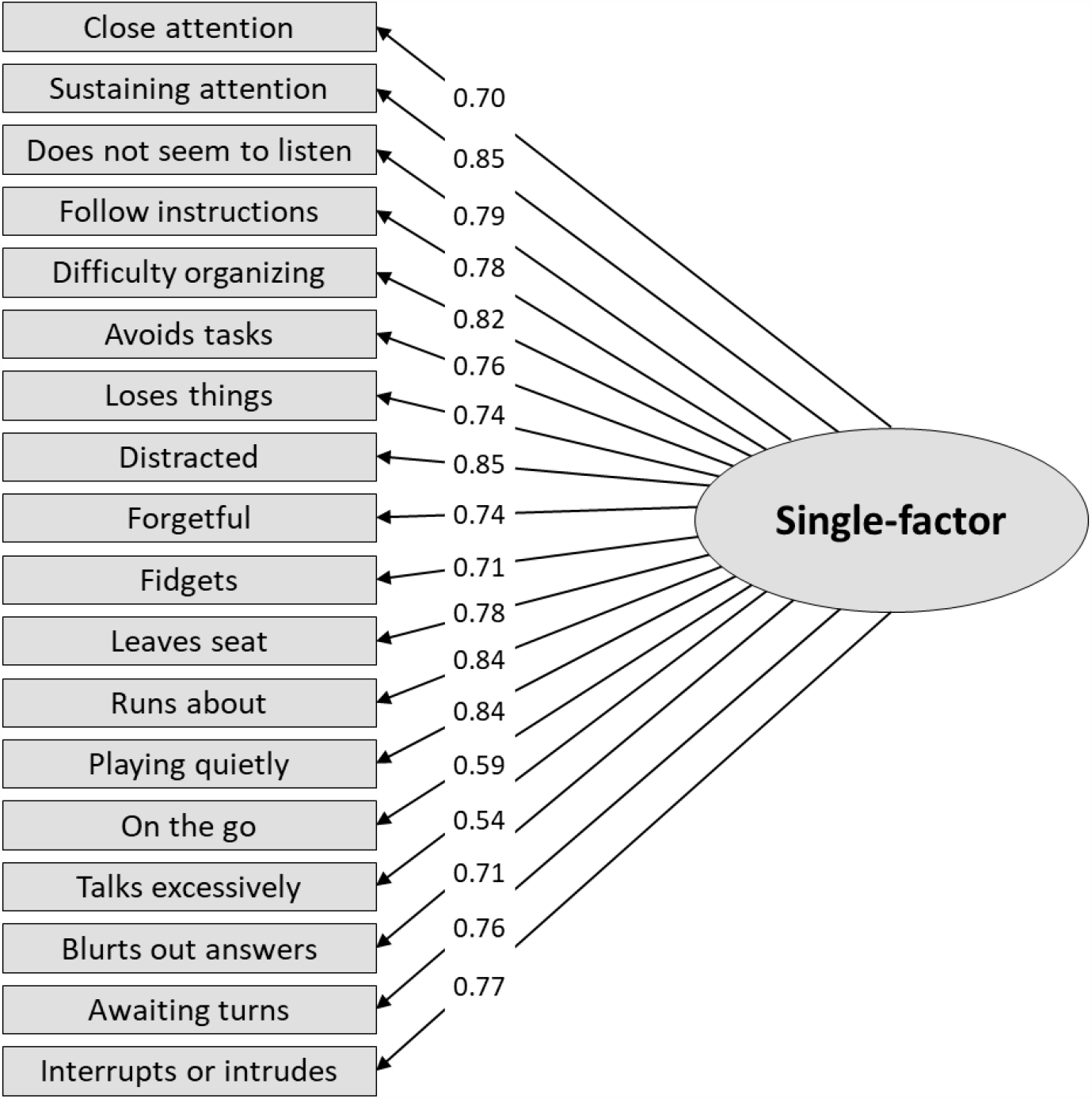
Single-factor model in the general population sample (N=2044) Notes: Dashed lines = non-significant factor loadings (p>0.05). Gray lines = Negative factor loadings.

**Figure S2.**
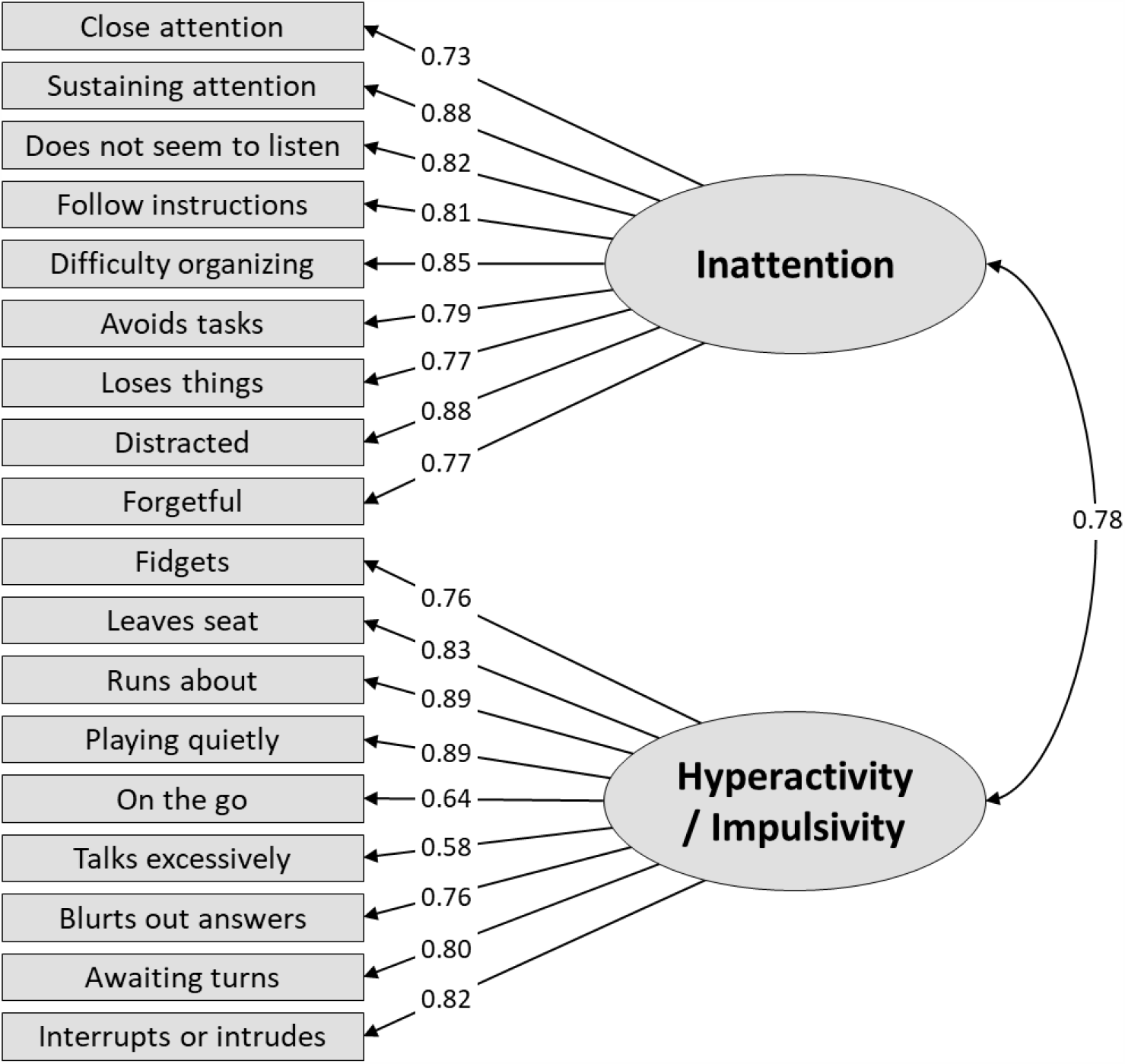
Correlated two-factor model in the general population sample (N=2044) Notes: Dashed lines = non-significant factor loadings (p>0.05). Gray lines = Negative factor loadings.

**Figure S3.**
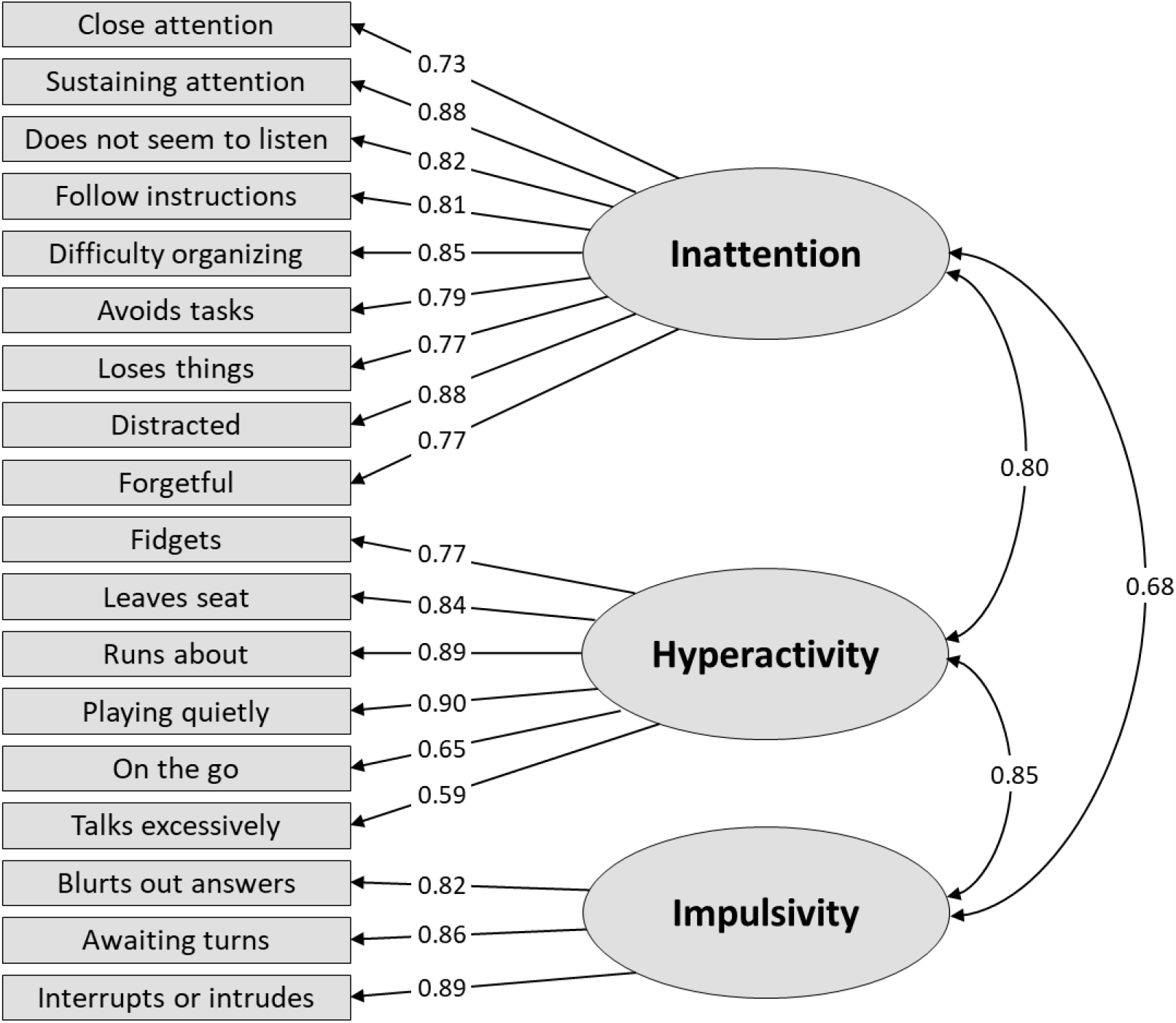
Correlated three-factor model with “talk excessively” as part of the Hyperactivity factor in the general population sample (N=2044) Notes: Dashed lines = non-significant factor loadings (p>0.05). Gray lines = Negative factor loadings.

**Figure S4.**
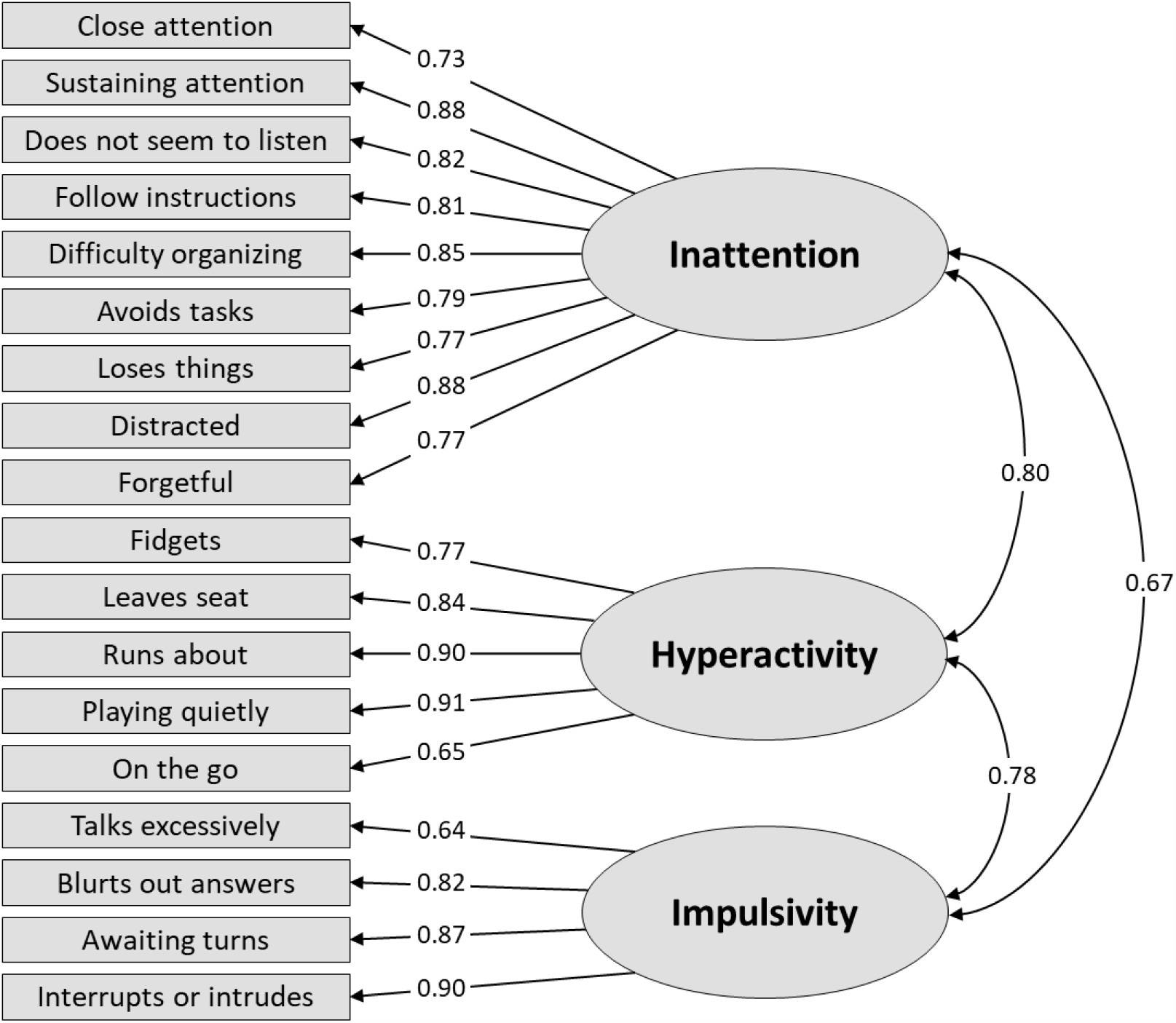
Correlated three-factor model with “talk excessively” as part of the Impulsivity factor in the general population sample (N=2044) Notes: Dashed lines = non-significant factor loadings (p>0.05). Gray lines = Negative factor loadings.

**Figure S5.**
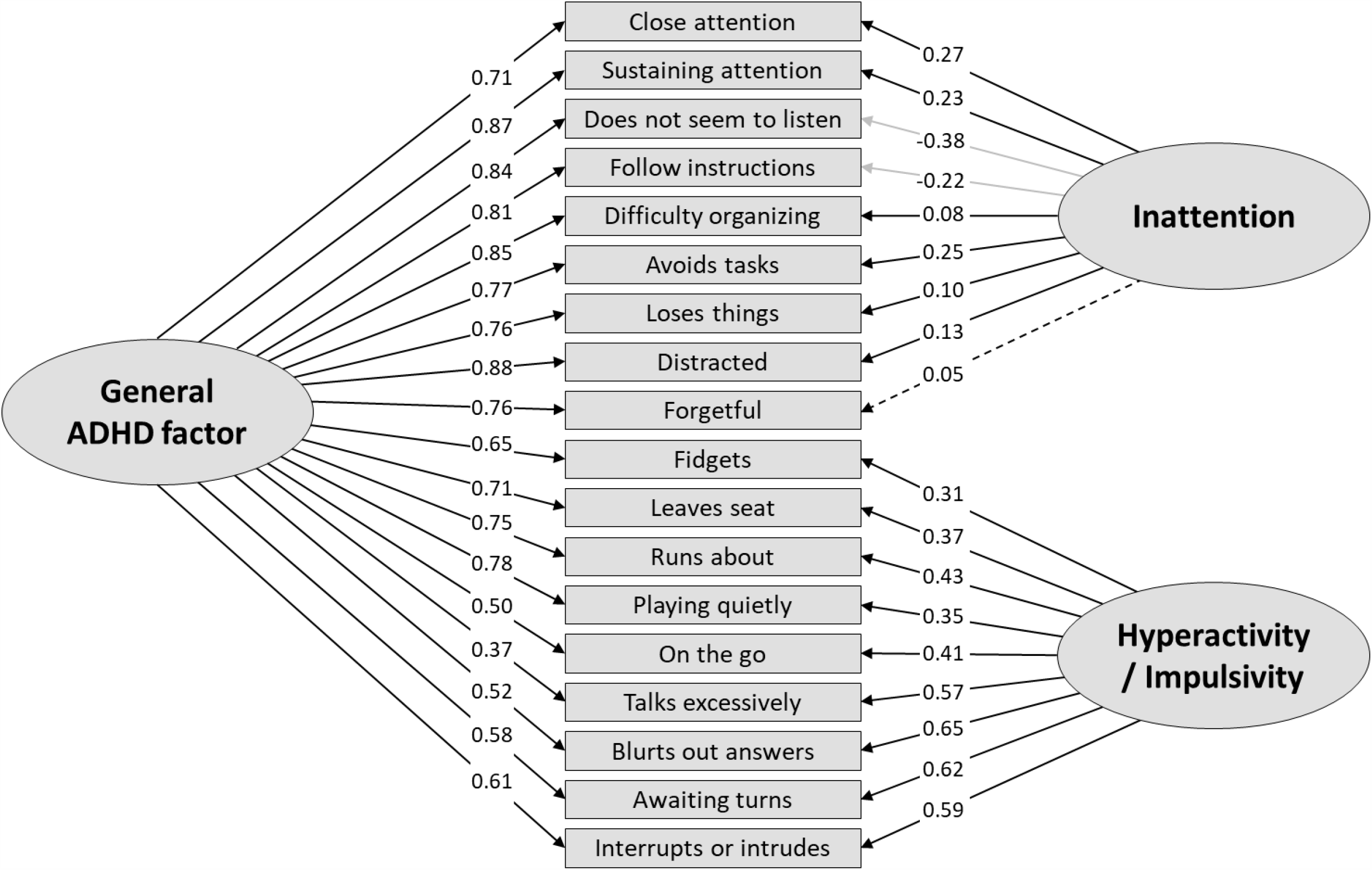
Bifactor model with two specific factors in the general population sample (N=2044) Notes: Dashed lines = non-significant factor loadings (p>0.05). Gray lines = Negative factor loadings.

**Figure S6.**
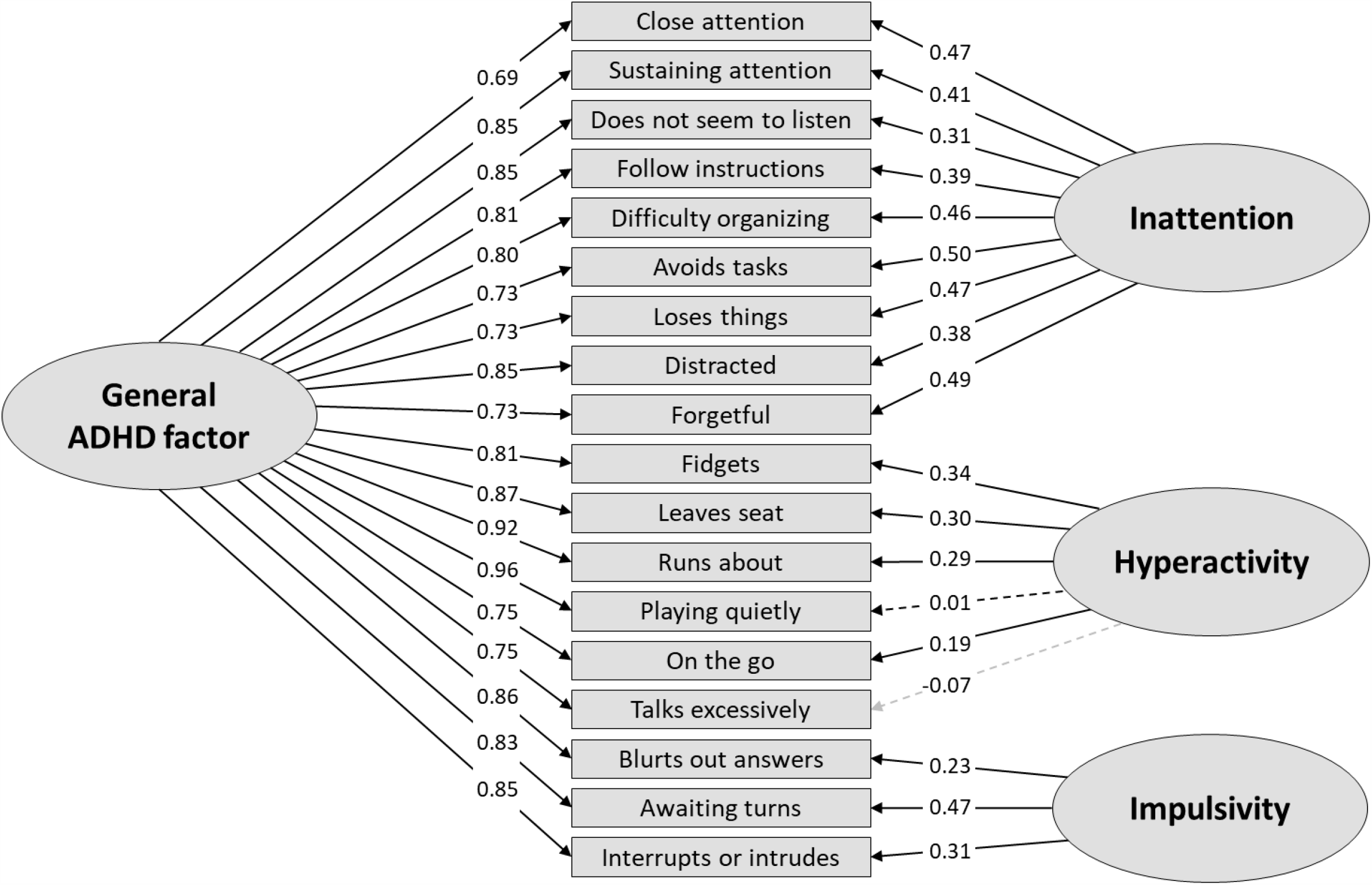
Bifactor model with three specific factors with “talk excessively” as part of the specific Hyperactivity factor in the general population sample (N=2044) Notes: Dashed lines = non-significant factor loadings (p>0.05). Gray lines = Negative factor loadings.

**Figure S7.**
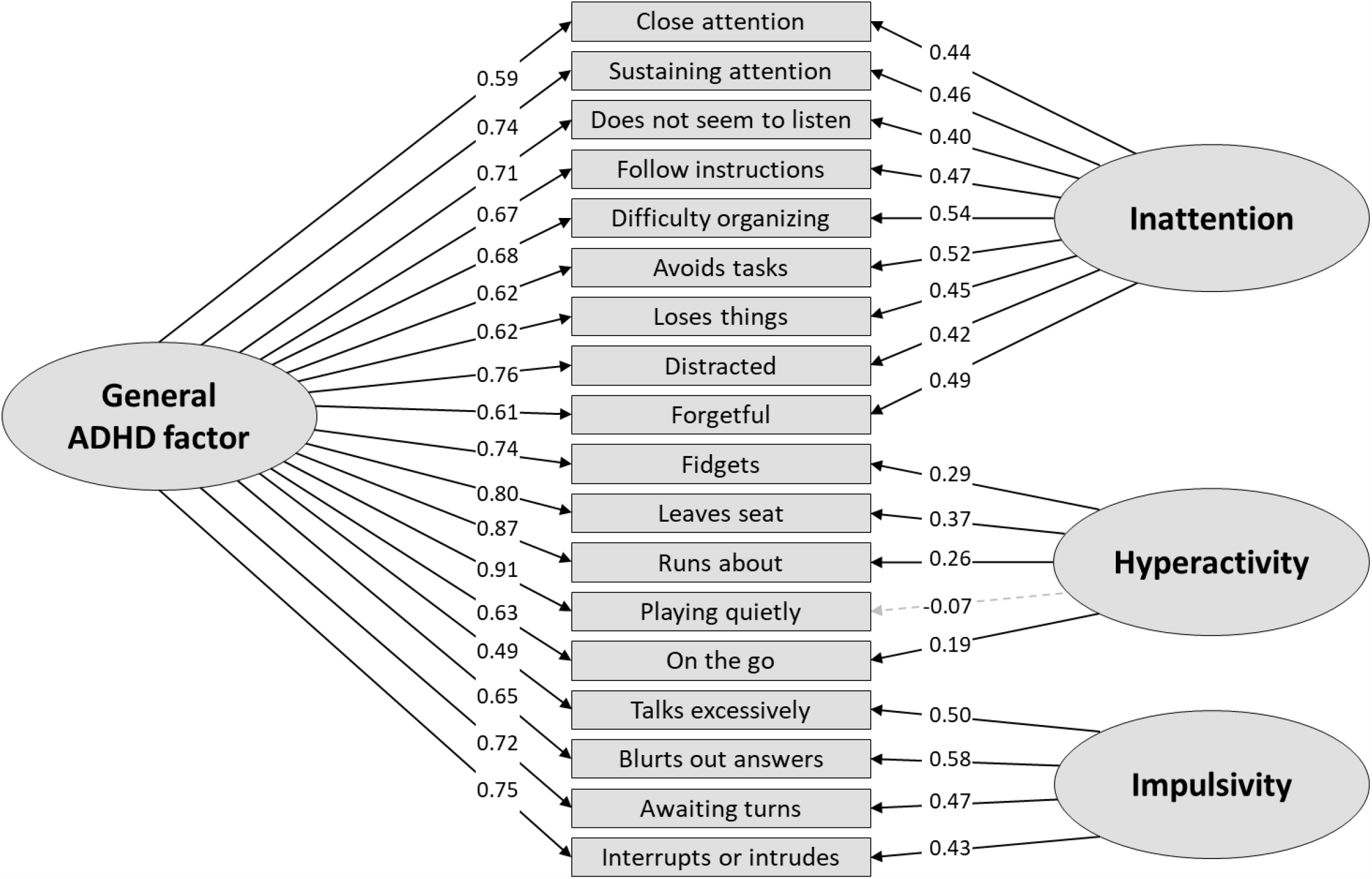
Bifactor model with three specific factors with “talk excessively” as part of the specific Impulsivity factor in the general population sample (N=2044) Notes: Dashed lines = non-significant factor loadings (p>0.05). Gray lines = Negative factor loadings.

**Figure S8.**
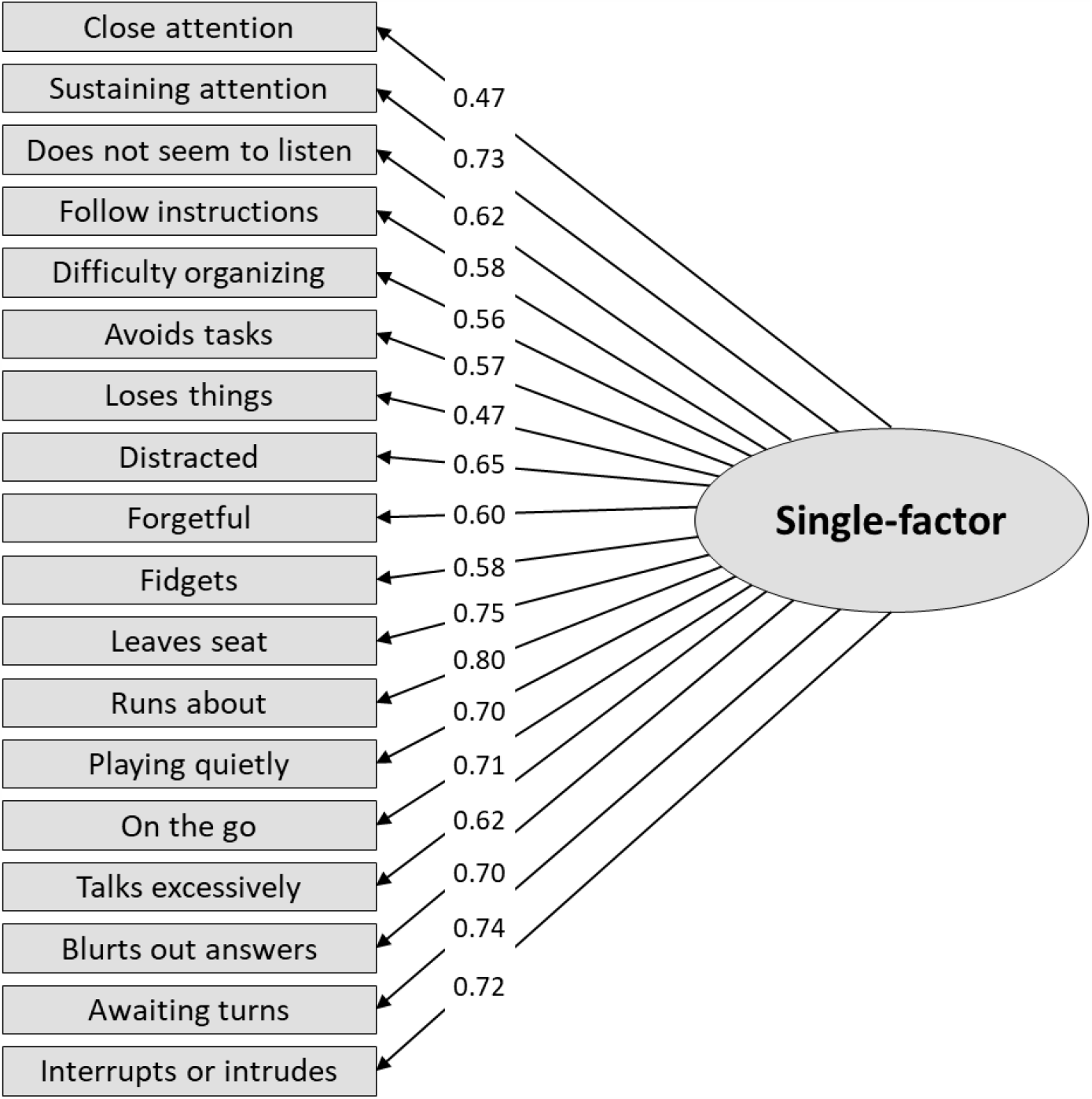
Single-factor model in the ADHD sample (N=147) Notes: Dashed lines = non-significant factor loadings (p>0.05). Gray lines = Negative factor loadings.

**Figure S9.**
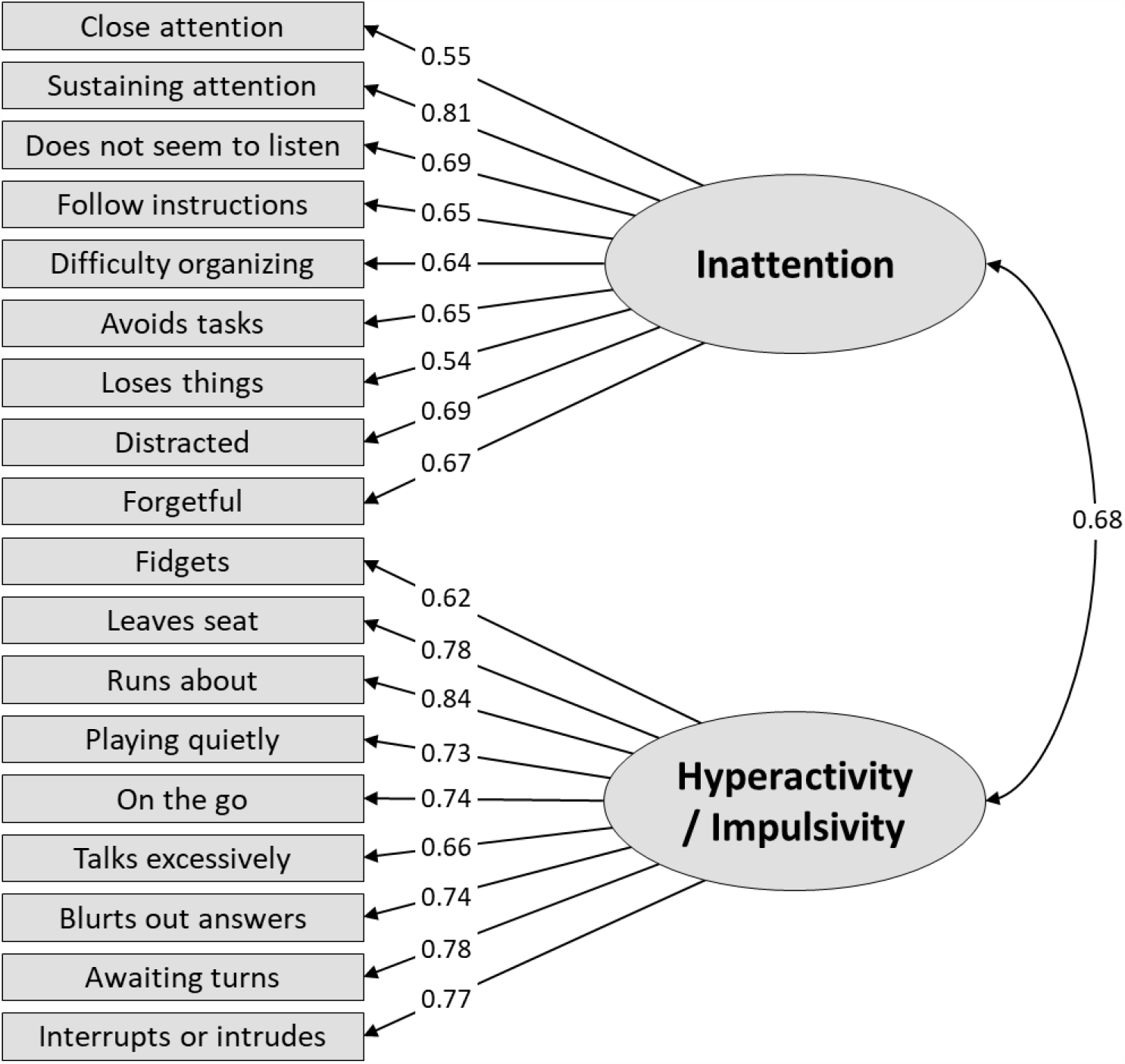
Correlated two-factor model in the ADHD sample (N=147) Notes: Dashed lines = non-significant factor loadings (p>0.05). Gray lines = Negative factor loadings.

**Figure S10.**
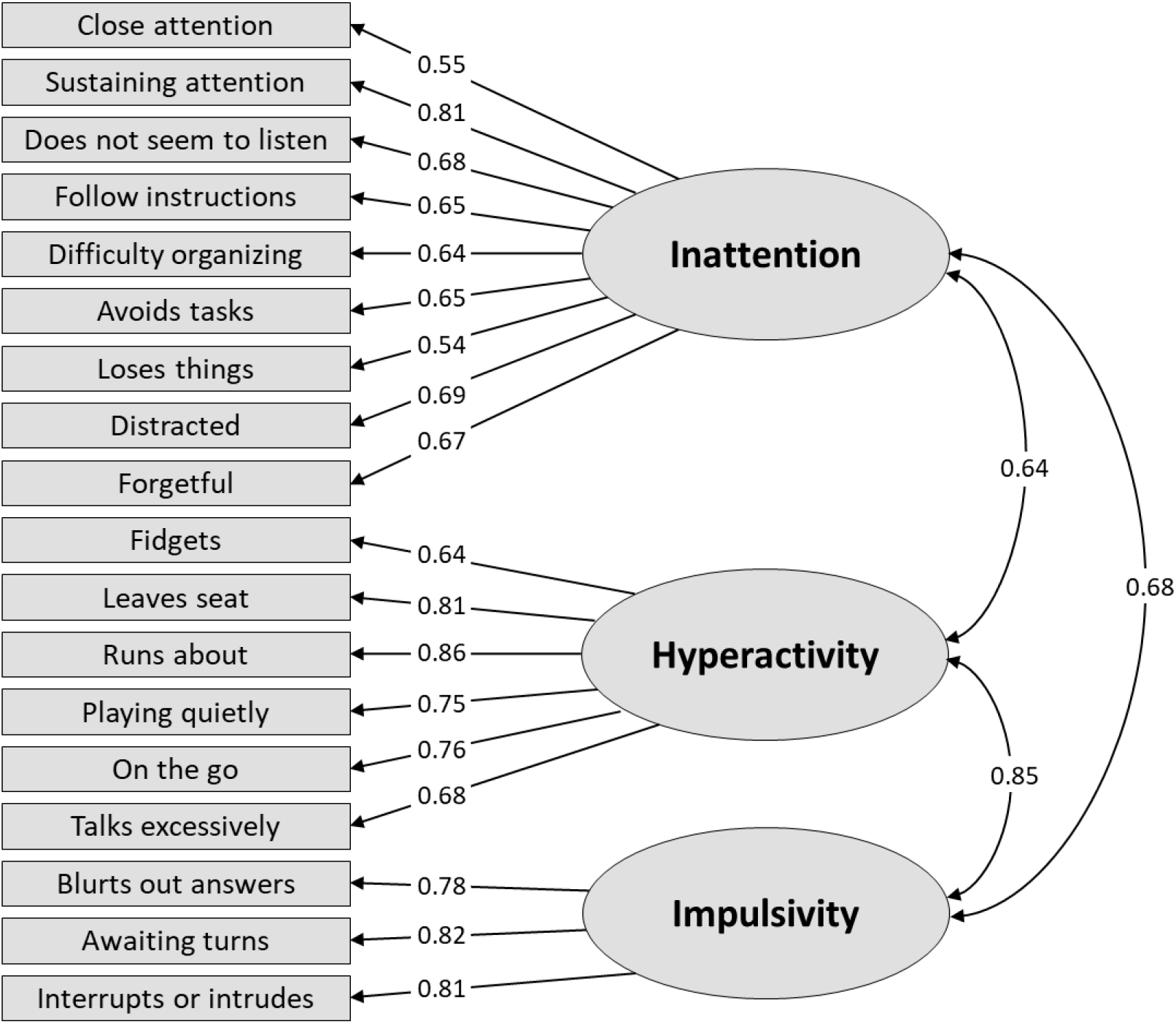
Correlated three-factor model with “talk excessively” as part of the Hyperactivity factor in the ADHD sample (N=147) Notes: Dashed lines = non-significant factor loadings (p>0.05). Gray lines = Negative factor loadings.

**Figure S11.**
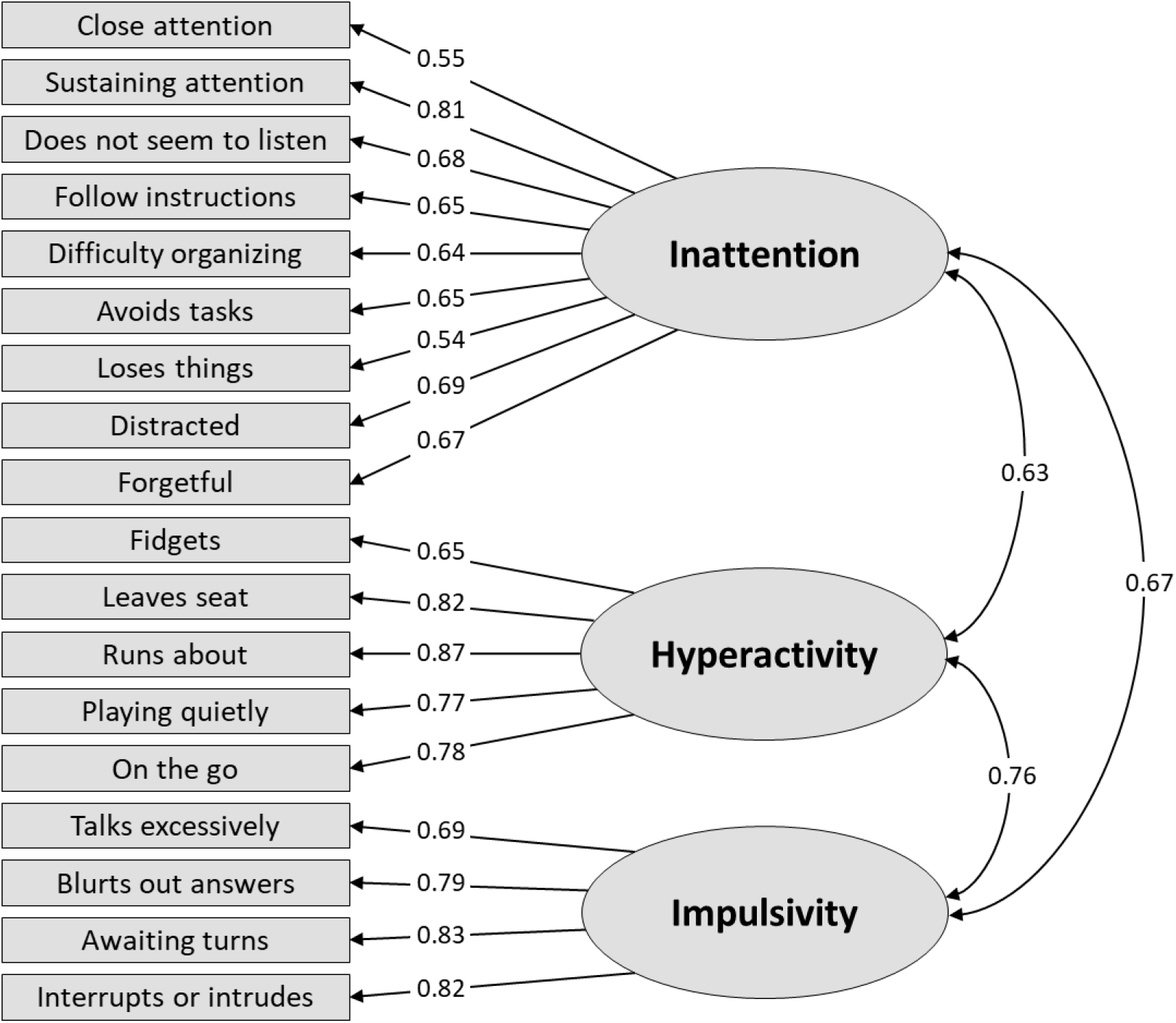
Correlated three-factor model with “talk excessively” as part of the Impulsivity factor in the ADHD sample (N=147) Notes: Dashed lines = non-significant factor loadings (p>0.05). Gray lines = Negative factor loadings.

**Figure S12.**
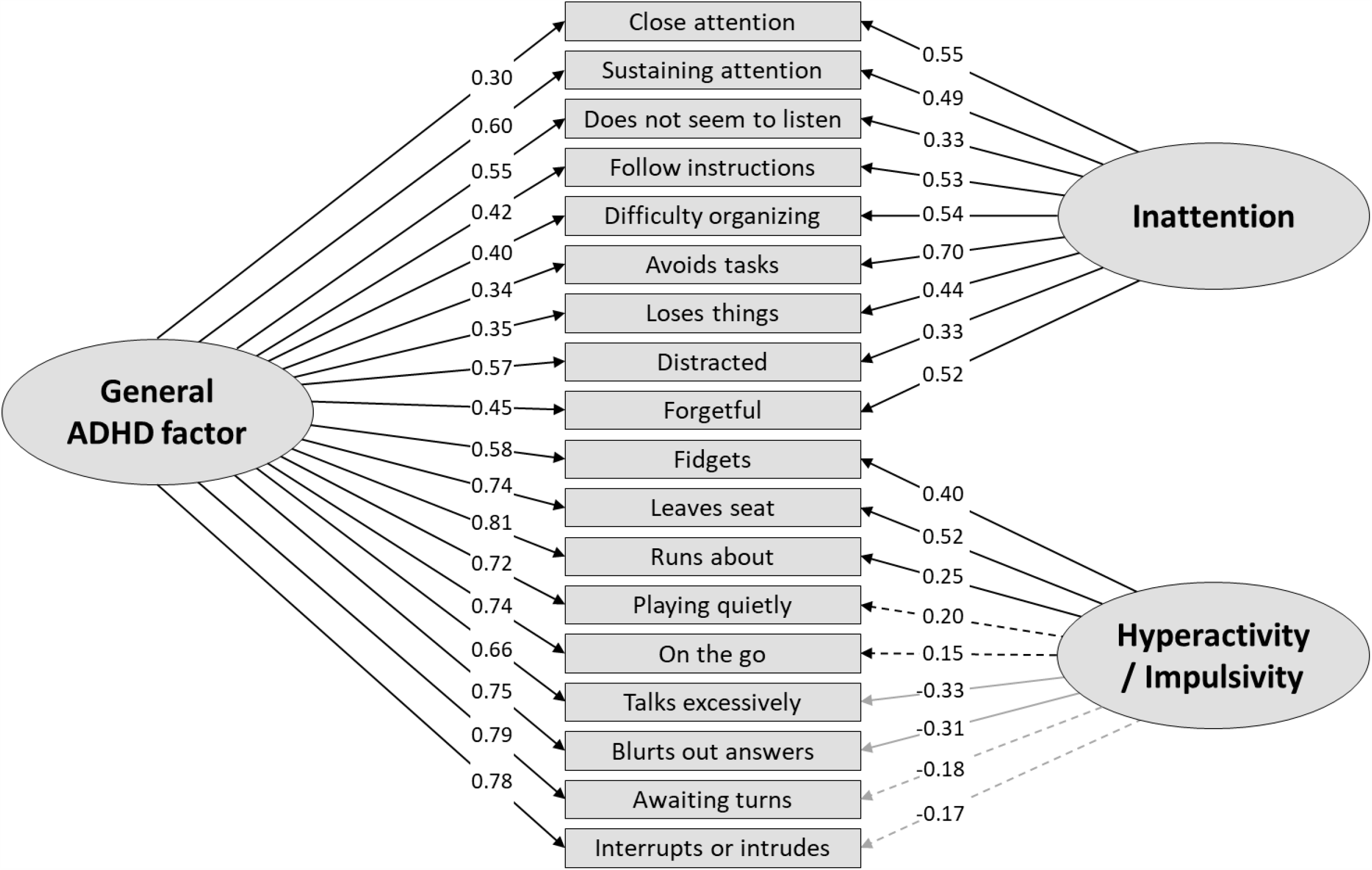
Bifactor model with two specific factors in the ADHD sample (N=147)

**Figure S13.**
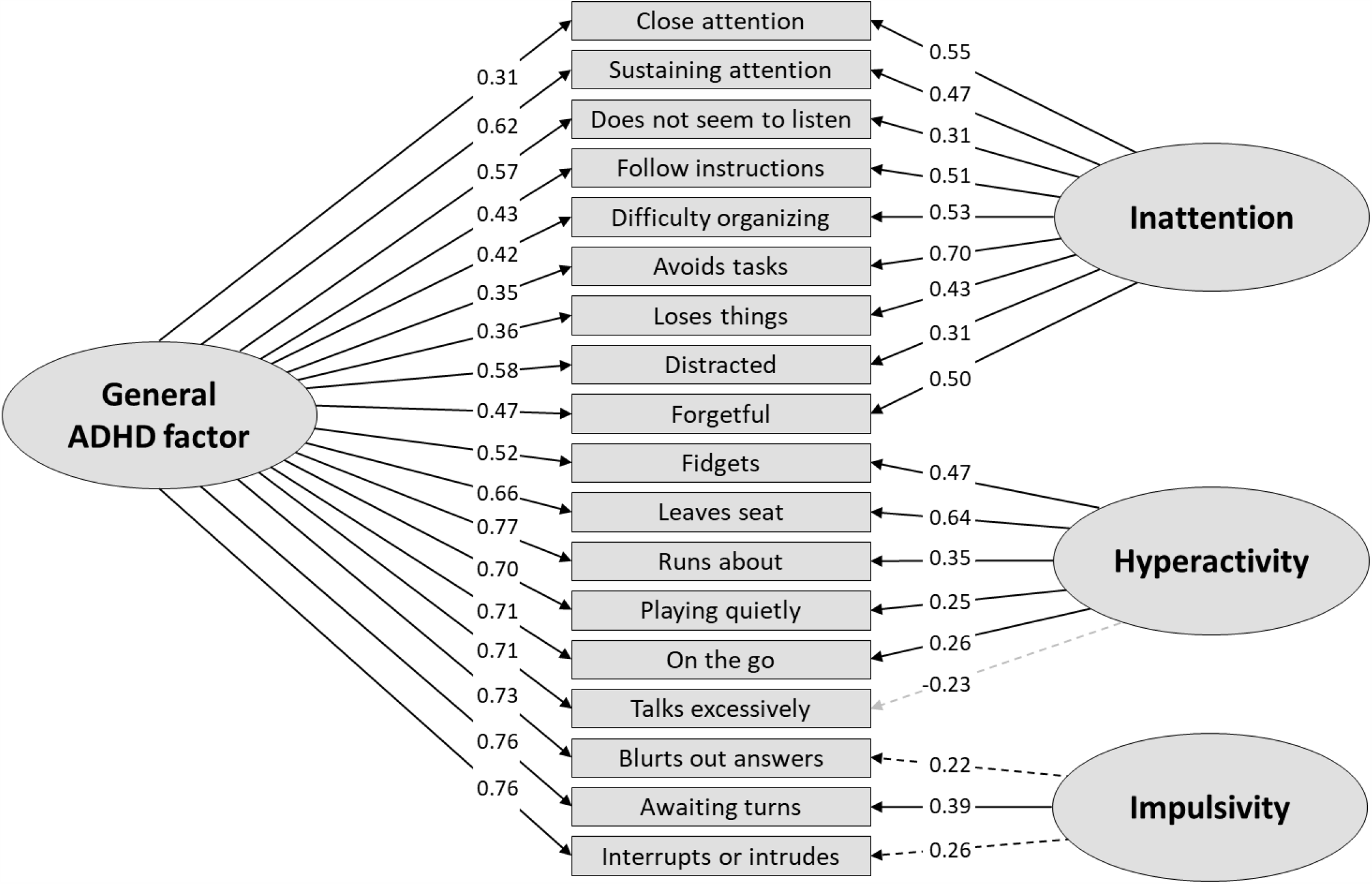
Bifactor model with three specific factors with “talk excessively” as part of the specific Hyperactivity factor in the ADHD sample (N=147) Notes: Dashed lines = non-significant factor loadings (p>0.05). Gray lines = Negative factor loadings.

**Figure S14.**
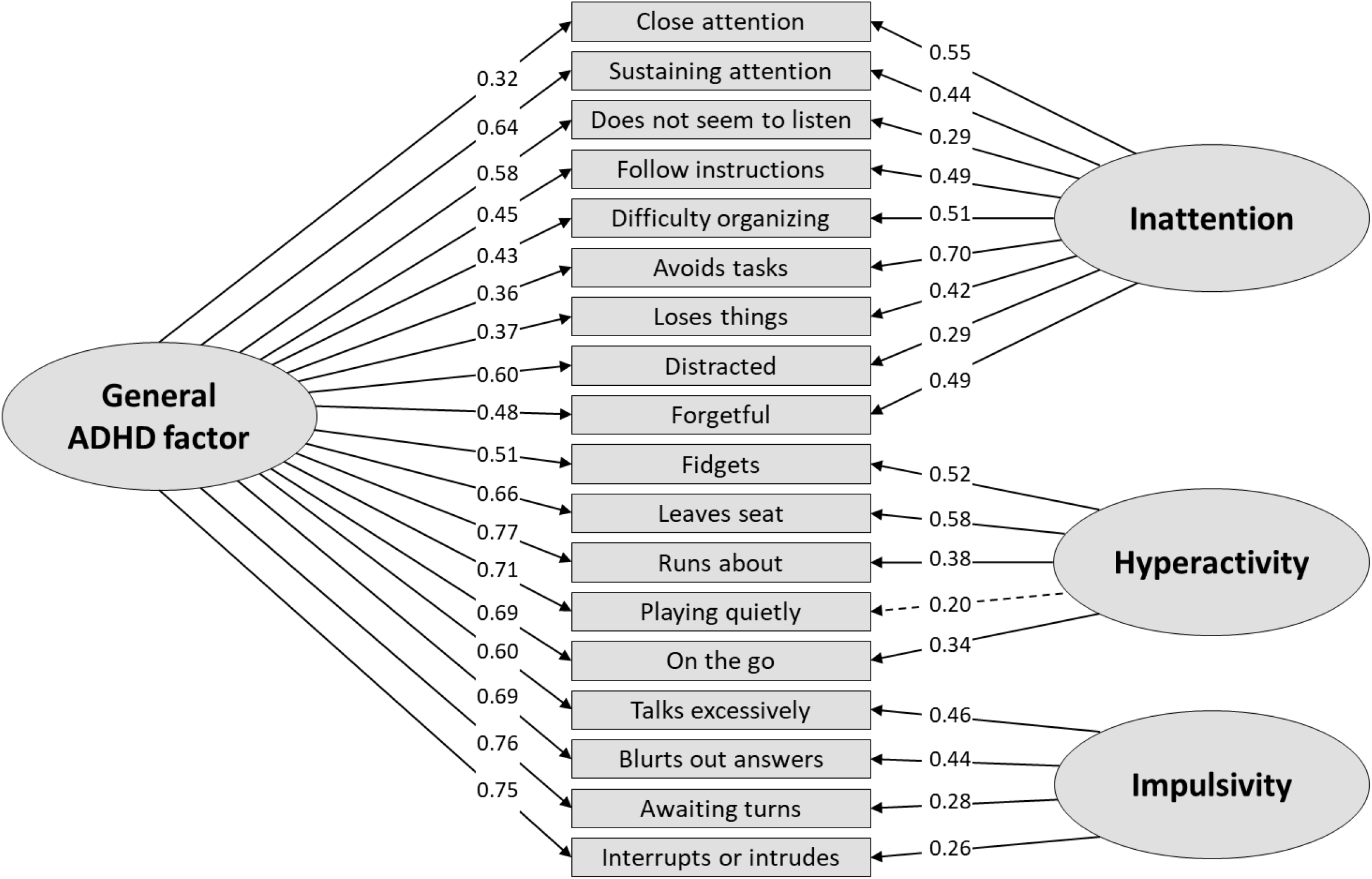
Bifactor model with three specific factors with “talk excessively” as part of the specific Impulsivity factor in the ADHD sample (N=147) Notes: Dashed lines = non-significant factor loadings (p>0.05). Gray lines = Negative factor loadings.

## Online Resource 3

**Table S1.** Gender-and-age-stratified ADHD-RS-IV scores in the ADHD sample

|  | **n** | **M** | **SD** | **Mdn** |
| --- | --- | --- | --- | --- |
| **Girls with ADHD aged 6-9** |  |  |  |  |
| Total score | 17 | 34.47 | 8.13 | 35 |
| Inattention | 17 | 18.59 | 5.11 | 20 |
| Hyperactivity/impulsivity | 17 | 15.94 | 5.60 | 17 |
| Hyperactivity alone<sup>a</sup> | 17 | 7.76 | 3.54 | 9 |
| Impulsivity alone<sup>b</sup> | 17 | 8.18 | 2.53 | 8 |
| **Girls with ADHD aged 10+** |  |  |  |  |
| Total score | 19 | 28.63 | 11.10 | 29 |
| Inattention | 19 | 17.05 | 5.28 | 18 |
| Hyperactivity/impulsivity | 19 | 11.53 | 7.12 | 10 |
| Hyperactivity alone<sup>a</sup> | 18 | 5.89 | 4.38 | 6.50 |
| Impulsivity alone<sup>b</sup> | 19 | 5.79 | 3.52 | 6 |
| **Boys with ADHD aged 6-9** |  |  |  |  |
| Total score | 65 | 30.80 | 10.87 | 30 |
| Inattention | 65 | 15.45 | 5.62 | 16 |
| Hyperactivity/impulsivity | 65 | 15.35 | 6.27 | 15 |
| Hyperactivity alone<sup>a</sup> | 65 | 8.75 | 3.87 | 9 |
| Impulsivity alone<sup>b</sup> | 65 | 6.60 | 3.01 | 6 |
| **Boys with ADHD aged 10+** |  |  |  |  |
| Total score | 64 | 30.38 | 9.39 | 30 |
| Inattention | 64 | 17.14 | 5.00 | 18 |
| Hyperactivity/impulsivity | 64 | 13.19 | 5.63 | 13 |
| Hyperactivity alone<sup>a</sup> | 64 | 7.27 | 3.43 | 7 |
| Impulsivity alone<sup>b</sup> | 64 | 5.94 | 2.87 | 6 |

Notes: n = sample size; M=Mean; SD=Standard deviation; Mdn=Median; ADHD-RS-IV= Attention-Deficit/Hyperactivity Disorder–Rating Scale IV. <sup>a</sup> This score is based on 5 items (i.e., “fidgets/squirms”, “leaves seat”, “runs about”, “difficulty playing quietly”, and “on the go”). <sup>b</sup> This score is based on 4 items (i.e., “talks excessively”, “blurts out answers”, “difficulty awaiting turns”, and “interrupts or intrudes”).

**Table S2.** Comparisons on summed ADHD-RS-IV scores between samples

|  | **ADHD<sup>a</sup>** | **General**<br>**population<sup>b</sup>** | **Mean difference**<br>**[95 % CI]** | ***t* test** |
| --- | --- | --- | --- | --- |
| **ADHD-RS-IV score, M (SD)** |  |  |  |  |
| Total score | 30.76 (10.10) | 10.09 (8.68) | 20.68 [19.08; 22.27] | *t*(184.08)=25.55, *p*<0.0001 |
| Inattention | 16.61 (5.35) | 5.30 (4.98) | 11.31 [10.52; 12.11] | *t*(2207)=27.90, *p*<0.0001 |
| Hyperactivity/impulsivity | 14.13 (6.18) | 4.79 (5.46) | 9.35 [8.38; 10.32] | *t*(178.03)=19.03, *p*<0.0001 |
| Hyperactivity alone<sup>c</sup> | 7.76 (3.81) | 2.20 (2.60) | 5.56 [4.96; 6.16] | *t*(175.37)=18.32, *p*<0.0001 |
| Impulsivity alone<sup>d</sup> | 6.41 (3.03) | 2.59 (2.34) | 3.82 [3.35; 4.30] | *t*(180.19)=15.84, *p*<0.0001 |

Notes: ADHD-RS-IV= Attention-Deficit/Hyperactivity Disorder–Rating Scale IV; CI = Confidence Interval; M=Mean; SD=Standard deviation.
<sup>a</sup> N=164–165. <sup>b</sup> N=2044. <sup>c</sup> Based on 5 items (i.e., “fidgets/squirms,” “leaves seat”, “runs about”, “difficulty playing quietly”, and “on the go”). <sup>d</sup> Based on 4 items (i.e., “talks excessively”, “blurts out answers”, “difficulty awaiting turns”, and “interrupts or intrudes”).

## Notes

### Clinical Trial

The study was not registered because this study is not a clinical trial but based on a parent-reported survey of ADHD symptoms in schoolchildren in the general population and data from another study of sleep problems in children with ADHD (which was not a clinical trial).

### Author Declarations

The survey was registered with and approved by the Danish Data Protection Agency (1-16-02-542-15), and the data was processed and stored in accordance with the European Union General Data Protection Regulation. Ethical review board approval is not required for survey-based studies in Denmark. The study by Virring et al. providing data for this analysis was approved by the Danish Regional Ethics Committee (M-20100231) and the Danish Data Protection Agency (2010-41-5472).

## REFERENCES

1. World Health Organization (1992) The ICD-10 classification of mental and behavioural disorders: clinical descriptions and diagnostic guidelines. World Health Organization, Geneva

2. American Psychiatric Association (2013) Diagnostic and statistical manual of mental disorders: DSM-5. American Psychiatric Association, 5th ed. Author, Washington, DC

3. Lange KW, Reichl S, Lange KM, et al (2010) The history of attention deficit hyperactivity disorder. ADHD Atten Deficit Hyperact Disord 2:241–255. https://doi.org/10.1007/s12402-010-0045-8

4. Alexandre JL, Lange AM, Bilenberg N, et al (2018) The ADHD rating scale-IV preschool version: Factor structure, reliability, validity, and standardisation in a Danish community sample. Res Dev Disabil. https://doi.org/10.1016/j.ridd.2018.05.006

5. McGoey KE, Schreiber J, Venesky L, et al (2015) Factor Structure of Attention Deficit Hyperactivity Disorder Symptoms for Children Age 3 to 5 Years. J Psychoeduc Assess 33:430–438. https://doi.org/10.1177/0734282914554255

6. Nichols JQVA, Shoulberg EK, Garner AA, et al (2017) Exploration of the Factor Structure of ADHD in Adolescence through Self, Parent, and Teacher Reports of Symptomatology. J Abnorm Child Psychol 45:625–641. https://doi.org/10.1007/s10802-016-0183-3

7. Pillow DR, Pelham WE, Hoza B, et al (1998) Confirmatory factor analyses examining attention deficit hyperactivity disorder symptoms and other childhood disruptive behaviors. J Abnorm Child Psychol 26:293–309. https://doi.org/10.1023/A:1022658618368

8. Bauermeister JJ, Canino G, Polanczyk G, Rohde LA (2010) ADHD Across Cultures: Is There Evidence for a Bidimensional Organization of Symptoms? J Clin Child Adolesc Psychol 39:362–372. https://doi.org/10.1080/15374411003691743

9. DuPaul GJ, Reid R, Anastopoulos AD, et al (2016) Parent and teacher ratings of attention-deficit/hyperactivity disorder symptoms: Factor structure and normative data. Psychol Assess 28:214–225. https://doi.org/10.1037/pas0000166

10. Gomez R, Harvey J, Quick C, et al (1999) DSM-IV AD/HD: Confirmatory Factor Models, Prevalence, and Gender and Age Differences Based on Parent and Teacher Ratings of Australian Primary School Children. J Child Psychol Psychiatry 40:265–274. https://doi.org/10.1017/S0021963098003321

11. Ryser GR, Campbell HL, Miller BK (2010) Confirmatory Factor Analysis of the Scales for Diagnosing Attention Deficit Hyperactivity Disorder (SCALES). Educ Psychol Meas 70:844–857. https://doi.org/10.1177/0013164410366696

12. Wolraich ML, Lambert EW, Baumgaertel A, et al (2003) Teachers’ screening for attention deficit/hyperactivity disorder: Comparing multinational samples on teacher ratings of ADHD. J Abnorm Child Psychol 31:445–455. https://doi.org/10.1023/A:1023847719796

13. Willcutt EG, Nigg JT, Pennington BF, et al (2012) Validity of DSM-IV attention deficit/hyperactivity disorder symptom dimensions and subtypes. J Abnorm Psychol 121:991–1010. https://doi.org/10.1037/a0027347

14. Holzinger KJ, Swineford F (1937) The Bi-factor method. Psychometrika 2:41–54. https://doi.org/10.1007/BF02287965

15. Sturm A, McCracken JT, Cai L (2019) Evaluating the Hierarchical Structure of ADHD Symptoms and Invariance Across Age and Gender. Assessment 26:508–523. https://doi.org/10.1177/1073191117714559

16. Arias VB, Ponce FP, Núñez DE (2018) Bifactor Models of Attention-Deficit/Hyperactivity Disorder (ADHD): An Evaluation of Three Necessary but Underused Psychometric Indexes. Assessment 25:885–897. https://doi.org/10.1177/1073191116679260

17. Reise SP (2012) The Rediscovery of Bifactor Measurement Models. Multivariate Behav Res 47:667–696. https://doi.org/10.1080/00273171.2012.715555

18. Toplak ME, Sorge GB, Flora DB, et al (2012) The hierarchical factor model of ADHD: invariant across age and national groupings? J Child Psychol Psychiatry 53:292–303. https://doi.org/10.1111/j.1469-7610.2011.02500.x

19. Toplak ME, Pitch A, Flora DB, et al (2009) The Unity and Diversity of Inattention and Hyperactivity/Impulsivity in ADHD: Evidence for a General Factor with Separable Dimensions. J Abnorm Child Psychol 37:1137–1150. https://doi.org/10.1007/s10802-009-9336-y

20. Caci HM, Morin AJ, Tran A (2016) Teacher Ratings of the ADHD-RS IV in a Community Sample: Results From the ChiP-ARD Study. J Atten Disord 20:434–444. https://doi.org/10.1177/1087054712473834

21. Arias VB, Ponce FP, Martínez-Molina A, et al (2016) General and specific attention-deficit/hyperactivity disorder factors of children 4 to 6 years of age: An exploratory structural equation modeling approach to assessing symptom multidimensionality. J Abnorm Psychol 125:125–137. https://doi.org/10.1037/abn0000115

22. Allan DM, Lonigan CJ (2019) Examination of the Structure and Measurement of Inattentive, Hyperactive, and Impulsive Behaviors from Preschool to Grade 4. J Abnorm Child Psychol 47:975–987. https://doi.org/10.1007/s10802-018-0491-x

23. Rodenacker K, Hautmann C, Görtz-Dorten A, Döpfner M (2016) Bifactor Models Show a Superior Model Fit: Examination of the Factorial Validity of Parent-Reported and Self-Reported Symptoms of Attention-Deficit/Hyperactivity Disorders in Children and Adolescents. Psychopathology 49:31–39. https://doi.org/10.1159/000442295

24. Gibbins C, Toplak ME, Flora DB, et al (2012) Evidence for a General Factor Model of ADHD in Adults. J Atten Disord 16:635–644. https://doi.org/10.1177/1087054711416310

25. Morin AJS, Tran A, Caci H (2016) Factorial Validity of the ADHD Adult Symptom Rating Scale in a French Community Sample: Results From the ChiP-ARD Study. J Atten Disord 20:530–541. https://doi.org/10.1177/1087054713488825

26. Willoughby MT, Fabiano GA, Schatz NK, et al (2019) Bifactor Models of Attention Deficit/Hyperactivity Symptomatology in Adolescents. Assessment 26:799–810. https://doi.org/10.1177/1073191117698755

27. Ogg JA, Bateman L, Dedrick RF, Suldo SM (2016) The Relationship Between Life Satisfaction and ADHD Symptoms in Middle School Students. J Atten Disord 20:390–399. https://doi.org/10.1177/1087054714521292

28. Dumenci L, Mcconaughy SH, Achenbach TM (2004) A Hierarchical Three-Factor Model of Inattention-Hyperactivity-Impulsivity Derived From the Attention Problems Syndrome of the Teacher’ s Report Form. Sch Psycology Rev 33:287–301

29. Gomez R, Vance A, Stavropoulos V (2018) Test-Retest Measurement Invariance of Clinic Referred Children’s ADHD Symptoms. J Psychopathol Behav Assess 40:194–205. https://doi.org/10.1007/s10862-017-9636-4

30. Martel MM, von Eye A, Nigg J (2012) Developmental differences in structure of attention-deficit/hyperactivity disorder (ADHD) between childhood and adulthood. Int J Behav Dev 36:279–292. https://doi.org/10.1177/0165025412444077

31. Willoughby MT, Blanton ZE, Family Life Project Investigators (2015) Replication and External Validation of a Bi-Factor Parameterization of Attention Deficit/Hyperactivity Symptomatology. J Clin Child Adolesc Psychol 44:68–79. https://doi.org/10.1080/15374416.2013.850702

32. Wagner F, Martel MM, Cogo-Moreira H, et al (2016) Attention-deficit/hyperactivity disorder dimensionality: the reliable ‘g’ and the elusive ‘s’ dimensions. Eur Child Adolesc Psychiatry 25:83–90. https://doi.org/10.1007/s00787-015-0709-1

33. Stanton K, Forbes MK, Zimmerman M (2018) Distinct dimensions defining the Adult ADHD Self-Report Scale: Implications for assessing inattentive and hyperactive/impulsive symptoms. Psychol Assess 30:1549–1559. https://doi.org/10.1037/pas0000604

34. Normand S, Flora DB, Toplak ME, Tannock R (2012) Evidence for a General ADHD Factor from a Longitudinal General School Population Study. J Abnorm Child Psychol 40:555–567. https://doi.org/10.1007/s10802-011-9584-5

35. Park JL, Silveira M, Elliott M, et al (2018) Confirmatory Factor Analysis of the Structure of Adult ADHD Symptoms. J Psychopathol Behav Assess 40:573–585. https://doi.org/10.1007/s10862-018-9698-y

36. Rodenacker K, Hautmann C, Görtz-Dorten A, Döpfner M (2017) The Factor Structure of ADHD – Different Models, Analyses and Informants in a Bifactor Framework. J Psychopathol Behav Assess 39:92–102. https://doi.org/10.1007/s10862-016-9565-7

37. Bonifay W, Cai L (2017) On the Complexity of Item Response Theory Models. Multivariate Behav Res 52:465–484. https://doi.org/10.1080/00273171.2017.1309262

38. Bonifay W, Lane SP, Reise SP (2017) Three Concerns With Applying a Bifactor Model as a Structure of Psychopathology. Clin Psychol Sci 5:184–186. https://doi.org/10.1177/2167702616657069

39. Reise SP, Kim DS, Mansolf M, Widaman KF (2016) Is the Bifactor Model a Better Model or Is It Just Better at Modeling Implausible Responses? Application of Iteratively Reweighted Least Squares to the Rosenberg Self-Esteem Scale. Multivariate Behav Res 51:818–838. https://doi.org/10.1080/00273171.2016.1243461

40. Virring A, Lambek R, Jennum PJ, et al (2017) Sleep Problems and Daily Functioning in Children With ADHD: An Investigation of the Role of Impairment, ADHD Presentations, and Psychiatric Comorbidity. J Atten Disord 21:731–740. https://doi.org/10.1177/1087054714542001

41. DuPaul GJ, Power TJ, Anastopoulos AD, Reid R (1998) ADHD Rating Scale—IV: Checklists, Norms, and Clinical Interpretation. The Guilford Press, New York

42. DuPaul GJ, Anastopoulos AD, Power TJ, et al (1998) Parent Ratings of Attention-Deficit/Hyperactivity Disorder Symptoms: Factor Structure and Normative Data. J Psychopathol Behav Assess 20:83–102. https://doi.org/10.1023/A:1023087410712

43. Szomlaiski N, Dyrborg J, Rasmussen H, et al (2008) Validity and clinical feasibility of the ADHD rating scale (ADHD-RS) A Danish Nationwide Multicenter Study. Acta Paediatr 98:397–402. https://doi.org/10.1111/j.1651-2227.2008.01025.x

44. Statistics Denmark. (2019) StatBank Denmark, Table FOLK1E: Population at the first day of the quarter by region, sex, age, and ancestry. https://www.statbank.dk/FOLK1E

45. Statistics Denmark. Statbank Denmark, Table INDKF111: Income for families by type of income, family type, unit, region and time. https://www.statbank.dk/INDKF111

46. Madsen KR, Damsgaard MT, Rubin M, et al (2016) Loneliness and Ethnic Composition of the School Class: A Nationally Random Sample of Adolescents. J Youth Adolesc 45:1350–1365. https://doi.org/10.1007/s10964-016-0432-3

47. Rich Madsen K, Trab Damsgaard M, Smith Jervelund S, et al (2016) Loneliness, immigration background and self-identified ethnicity: a nationally representative study of adolescents in Denmark. J Ethn Migr Stud 42:1977–1995. https://doi.org/10.1080/1369183X.2015.1137754

48. Statistics Denmark (2017) Documentation of statistics for Immigrants and Descendants 2017 Month 01. Statistics Denmark, København Ø, Denmark

49. Goodman R, Ford T, Richards H, et al (2000) The Development and Well-Being Assessment: Description and Initial Validation of an Integrated Assessment of Child and Adolescent Psychopathology. J Child Psychol Psychiatry 41:645–655. https://doi.org/10.1111/j.1469-7610.2000.tb02345.x

50. Kline P (1994) An easy guide to factor analysis. Routledge, London

51. Bentler PM (1990) Comparative fit indexes in structural models. Psychol Bull 107:238–246. https://doi.org/10.1037/0033-2909.107.2.238

52. Tucker LR, Lewis C (1973) A reliability coefficient for maximum likelihood factor analysis. Psychometrika 38:1–10. https://doi.org/10.1007/BF02291170

53. Steiger JH (1990) Structural Model Evaluation and Modification: An Interval Estimation Approach. Multivariate Behav Res 25:173–180. https://doi.org/10.1207/s15327906mbr2502_4

54. Hu L, Bentler PM (1999) Cutoff criteria for fit indexes in covariance structure analysis: Conventional criteria versus new alternatives. Struct Equ Model A Multidiscip J 6:1–55. https://doi.org/10.1080/10705519909540118

55. Hu L, Bentler PM (1998) Fit indices in covariance structure modeling: Sensitivity to underparameterized model misspecification. Psychol Methods 3:424–453. https://doi.org/10.1037/1082-989X.3.4.424

56. Ten Berge JMF, Sočan G (2004) The greatest lower bound to the reliability of a test and the hypothesis of unidimensionality. Psychometrika 69:613–625. https://doi.org/10.1007/BF02289858

57. Reise SP, Scheines R, Widaman KF, Haviland MG (2013) Multidimensionality and Structural Coefficient Bias in Structural Equation Modeling: A Bifactor Perspective. Educ Psychol Meas 73:5–26. https://doi.org/10.1177/0013164412449831

58. McDonald RP (1999) Test theory: A unified treatment. Lawrence Erlbaum, Hillsdale, NJ

59. Bentler PM (2009) Alpha, Dimension-Free, and Model-Based Internal Consistency Reliability. Psychometrika 74:137–143. https://doi.org/10.1007/s11336-008-9100-1

60. Reise SP, Bonifay WE, Haviland MG (2013) Scoring and Modeling Psychological Measures in the Presence of Multidimensionality. J Pers Assess 95:129–140. https://doi.org/10.1080/00223891.2012.725437

61. Gagne P, Hancock GR (2006) Measurement Model Quality, Sample Size, and Solution Propriety in Confirmatory Factor Models. Multivariate Behav Res 41:65–83. https://doi.org/10.1207/s15327906mbr4101_5

62. Hancock GR, Mueller RO (2001) Rethinking construct reliability within latent variable systems. In: Cudeck R, du Toit S, Sorbom D (eds) Structural equation modeling: Present and future—A Festschrift in honor of Karl Joreskog. Scientific Software, Lincolnwood, IL, pp 195–216

63. Rodriguez A, Reise SP, Haviland MG (2016) Applying Bifactor Statistical Indices in the Evaluation of Psychological Measures. J Pers Assess 98:223–237. https://doi.org/10.1080/00223891.2015.1089249

64. Gignac GE, Watkins MW (2013) Bifactor Modeling and the Estimation of Model-Based Reliability in the WAIS-IV. Multivariate Behav Res 48:639–662. https://doi.org/10.1080/00273171.2013.804398

65. R Core Team (2019) R: A language and environment for statistical computing. R Foundation for Statistical Computing, Vienna, Austria. http://www.r-project.org/

66. Rosseel Y (2012) lavaan: An R Package for Structural Equation Modeling. J Stat Softw 48:1–36. https://doi.org/10.18637/jss.v048.i02

67. Dueber DM (2019) Bifactor Indices Calculator (Version 0.1.0). https://cran.rproject.org/web/packages/BifactorIndicesCalculator/BifactorIndicesCalculator.pdf

68. StataCorp. (2019) Stata Statistical Software: Release 16. College Station, TX

69. Ullebø AK, Breivik K, Gillberg C, et al (2012) The factor structure of ADHD in a general population of primary school children. J Child Psychol Psychiatry Allied Discip 53:927–936. https://doi.org/10.1111/j.1469-7610.2012.02549.x

70. Smith LC, Tamm L, Hughes CW, Bernstein IH (2013) Separate and overlapping relationships of inattention and hyperactivity/impulsivity in children and adolescents with attention-deficit/hyperactivity disorder. ADHD Atten Deficit Hyperact Disord 5:9–20. https://doi.org/10.1007/s12402-012-0091-5

71. Martel MM, Von Eye A, Nigg JT (2010) Revisiting the latent structure of ADHD: is there a ‘g’ factor? J Child Psychol Psychiatry 51:905–914. https://doi.org/10.1111/j.1469-7610.2010.02232.x

72. Goh PK, Lee CA, Bansal PS, et al (2019) Interpretability and Validity of a Bifactor Model of ADHD in Young Adults: Assessing the General “g” and Specific IA and HI Factors. J Psychopathol Behav Assess. https://doi.org/10.1007/s10862-019-09774-7

73. Pelham WE, Evans SW, Gnagy EM, Greenslade KE (1992) Teacher ratings of DSM-III—R symptoms for the disruptive behavior disorders: Prevalence, factor analyses, and conditional probabilities in a special education sample. School Psych Rev 21:285–299

74. Statistics Denmark. Statbank Denmark, Table HFUDD10: Educational attainment (15-69 years) by region, ancestry, highest education completed, age and sex. www.statbank.dk/HFUDD10. Accessed 17 Mar 2020

